# Simulation-based Medical Emergency Training for Deep-Space Missions

**DOI:** 10.1101/2025.10.15.25338063

**Authors:** Susana Dias Alves, Mariana Peyroteo, Joana Godinho, João Lousada, Gonçalo Torrinha, Luís Velez Lapão

## Abstract

The Artemis program aims to establish the Lunar Gateway for missions beyond low Earth orbit, requiring astronauts to have greater medical independence. Earth Independent Medical Operations is the transition to autonomous astronaut care, necessitating proper equipment for managing medical emergencies. Analog missions simulate space missions on Earth, enabling safe and cost-effective testing of technologies, refining protocols, and enhancing decision-making skills. High-stress, realistic simulations are among the most effective training methods.

A new training system for managing medical emergencies during deep-space missions was developed, simulating real-time scenarios akin to a Situation Room. This system dynamically updates vital signs and critical data based on crew and mission control decisions through multiple-choice responses, assessing medical knowledge, procedural execution, and non-technical skills such as situational awareness, teamwork, task load, and communication.

An analog lunar mission simulation involving four astronauts and four mission control personnel served as the initial test. All astronauts underwent the same medical training except for the crew medical officer, who had relevant expertise. The scenario, focusing on a cerebrovascular accident, lasted about two hours, while supervisors evaluated teamwork using the Team Emergency Assessment Measure survey. Participants then completed NASA’s Task Load Index to assess their workload. Medical knowledge and procedural execution were measured through a multiple-choice questionnaire, with future research planned to include stress-related biomedical data.

In this simulation, Mission Control’s medical knowledge averaged 63.67%, while astronauts scored 50%; procedural knowledge was 51.3% and 56.82%, respectively. Teamwork assessments revealed strengths in cooperation but weaknesses in leadership and communication under time pressure. Task-load results indicated higher temporal demands for Mission control and greater frustration for astronauts, highlighting differences in workload perception across roles.

These findings suggest that structured simulations can identify gaps in both technical and non-technical skills, offering valuable improvement opportunities. The CHIRON platform proved effective as a training tool for acute medical emergencies in deep space and has potential for Earth-based crisis training, such as during emergencies or disaster response.

## 1. Introduction

For more than half a century, humanity has pursued exploration beyond Earth’s bounds—initially to the lunar surface during the Apollo program and now with ambitions set on Mars. The National Aeronautics and Space Administration (NASA) led the Artemis program, and the forthcoming Lunar Gateway will extend operations well beyond low-Earth orbit (LEO) [1], requiring astronauts to achieve unprecedented medical self-sufficiency. Deep-space missions face constraints such as communication delays, limited resupply, and the absence of rapid evacuation, which demands the transition to Earth-Independent Medical Operations (EIMO), defined as “*the transition of medical primacy from terrestrial to space-based assets to enable astronaut health and performance*” [2].

A medical emergency is an acute injury or illness that poses an immediate threat to life or long-term health [3]— it represents one of the most significant hazards in human spaceflight. NASA’s space medicine program, therefore, prioritizes sustaining life, minimizing health risks, and preventing injury in the unique environment of space[4]. Even seemingly minor conditions can compromise crew safety and mission objectives [5], [6], making structured emergency management essential. According to the Federal Emergency Management Agency (FEMA)[7], emergencies involve four key phases: mitigation, preparedness, response, and recovery. In space, this process is further complicated by distance, isolation, and limited medical capabilities[2]. For instance, a stroke, or cerebrovascular accident (CVA), is a medical emergency caused by a blocked blood vessel (ischemic) or bleeding in the brain (hemorrhagic)[8]. Immediate management is crucial to prevent permanent disability or death [9], compromising the mission itself [5].

Managing medical conditions in these missions will then demand advanced decision-support systems, robust and validated response protocols, and immersive, simulation-based training that faithfully replicates the rigors of deep-space medical contingencies [10], [11], [12].

### 1.1 Realistic Simulations for Training

For over 60 years, simulation-based training has been used in medicine and public health to prepare professionals for critical situations under controlled conditions. Research indicates that when simulations incorporate realistic complexity and stress, they are particularly effective in skill development [13].

The European Centre for Disease Prevention and Control has developed various types of simulation exercises for public health settings across the European Union. The most complex of these is the full-scale exercise, a scenario-based simulation designed to evaluate the effectiveness of emergency preparedness and response procedures and systems in real-world situations [14].

One example is a simulation run in a Situation Room (SR), which traditionally serves as the primary center for gathering intelligence, analysis, and coordination during a crisis. In SR-based simulations, participants receive continuous information updates and must make decisions that influence subsequent events, while supervisors evaluate both technical and non-technical skills. This approach allows crews to experience the time-critical pressures of real emergencies while practicing communication hierarchies and leadership [15].

### 1.2 Context of this research

To prepare astronauts and Mission Control Centre (MCC) personnel for the immediate management of medical emergencies in deep-space missions, the CHIRON (Crew Health OptImisation for em-eRgencies in deep-space missiONs) platform was developed within a situation room simulation-based training system. For this study, a simulation involving a CVA under lunar gateway mission conditions was created. With this system, it is possible to train and assess both technical skills, such as medical and procedural knowledge, and non-technical skills, including teamwork, task load, and communication.

### 2. Materials and methods

This project followed the Design Science and Research Methodology (DSRM), a scientific approach that focuses on designing artifacts to solve real-world problems [16][17]. DSRM is an iterative process that includes six steps: problem identification and motivation, defining objectives for a solution, design and development, demonstration, evaluation, and communication (Figure 1) [18].

**Figure 1.**
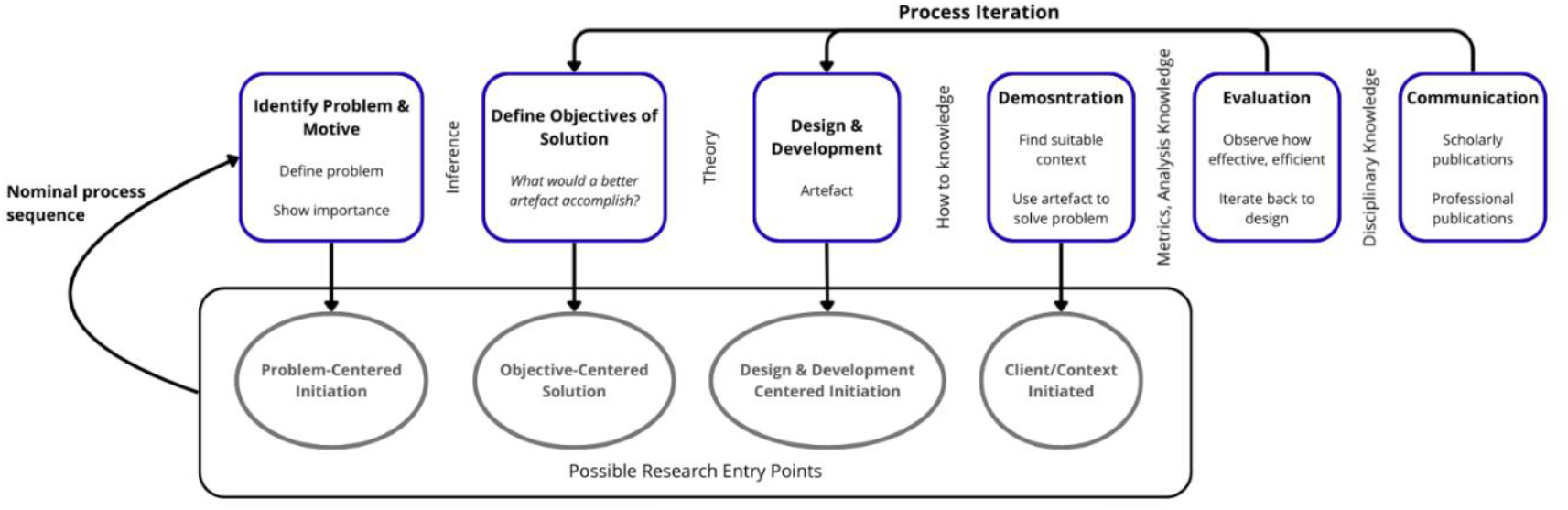
Design Science and Research Methodology process Adapted from Peffers et al. 2007[18]

**Figure 2.**
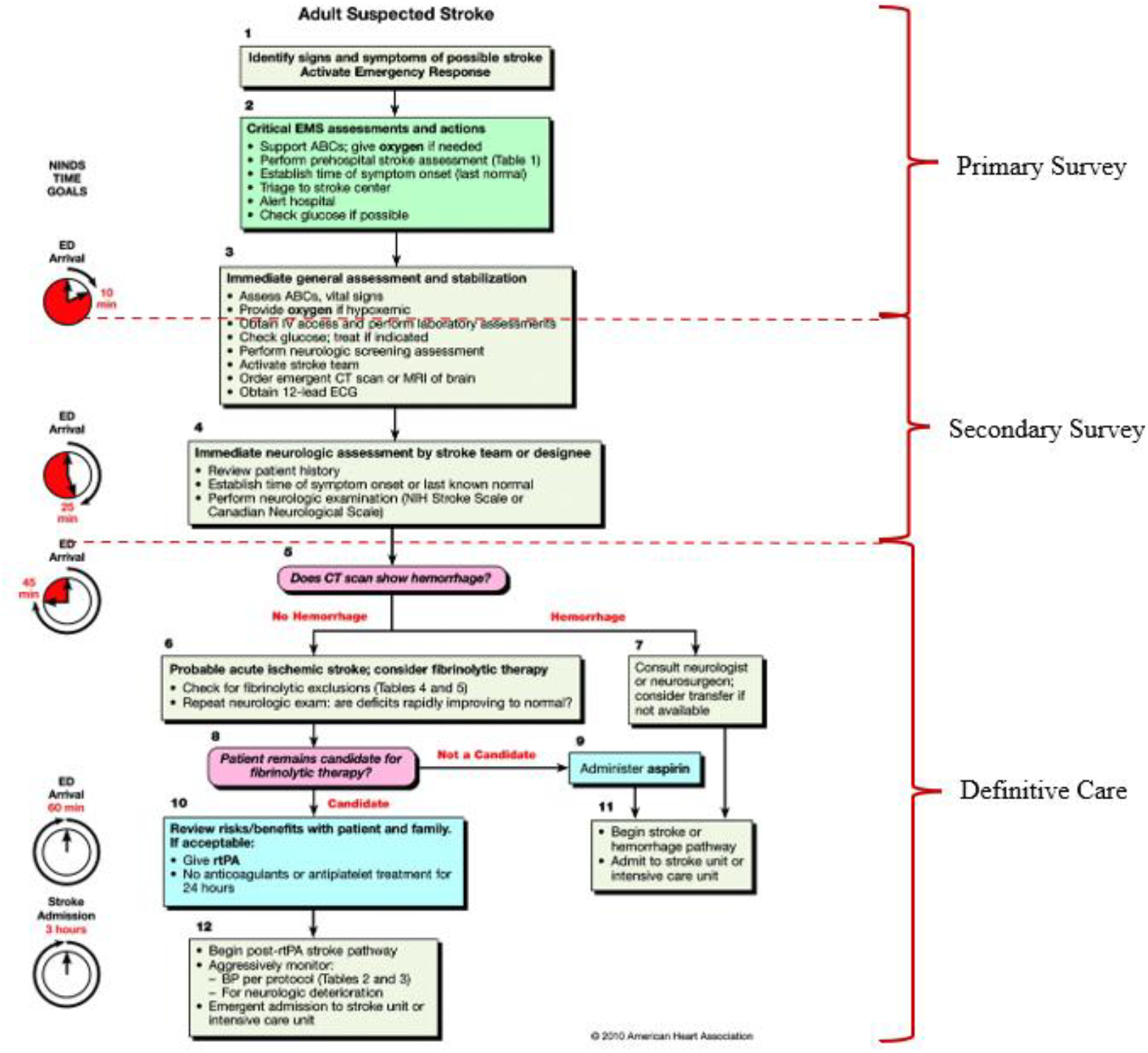
Separation of the steps of the Adult Suspected Stroke Algorithm[50] into the phases of emergency medical

### 2.1 Objectives of a Solution

Rosenberg et al. (2025) [19] highlight the need for in-flight protocols for quick diagnosis and treatment of emergency neurologic events, specifically CVAs, in missions beyond LEO. To date, there has been limited research examining team performance or the decision-making process during crises. These findings highlight the importance of training crews for CVA scenarios and adapting protocols for spaceflight, as well as the need for operational testing to verify their effectiveness.

In response, the CHIRON platform was developed, aiming at accomplishing three objectives:

- Reproduce deep-space conditions, including communication delays, mission abort scenarios, resource limitations, and physical and psychological distance.
- Apply existing ischemic stroke protocols in a spaceflight context.
- Strengthen personnel competencies during the response phase, focusing on medical and procedural skills, teamwork, and task load management.

### 2.2 Design and Development

The design process combined the storyline and the informatics system. The storyline was based on “*Handbook on Simulation Exercises in EU Public Health Settings*”[14] and the World Health Organization (WHO) “*Simulation Exercise Manual*”[20] and adapted to space conditions. Input was collected from the European Space Agency (ESA) experts (Flight Surgeon and Flight Director), scientific publications, and information retrieved from official websites. The final storyline reproduced CVA during extravehicular activity (EVA), with sequential injects of new information, decision points, and evolving physiological parameters.

Technically, CHIRON was implemented as a web-based application. The frontend, developed in Streamlit (version 1.47.0)[21], displayed vital signs, dashboards, and decision prompts. Supabase[22] powered the backend, providing authentication, database management, and real-time data handling. Communication between astronauts and MCC was conducted through two systems: Zoom (version 6.5.7)[23] and Asclepios COMMS.

### 2.3 Demonstration

The demonstration was conducted using a simulated SR scenario of a CVA occurring during an EVA in a Lunar Gateway mission. Two distinct crews participated: the astronaut crew and the MCC crew, each composed of four members. The astronaut crew included a Commander and three Flight Engineers (FEs). In this simulation, the Commander also served as the Crew Medical Officer (CMO) and Intravehicular 2 (IV2). FE-1 also acted as Extravehicular 1 (EV1), FE-2 as Intravehicular 1 (IV1), and FE-3 as Extravehicular 2 (EV2). On the MCC side, the team consisted of a Flight Director (FD), Flight Surgeon (FS), Biomedical Engineer (BME), and Capsule Communicator (CAPCOM).

As two iterations of the DSRM were conducted, two distinguished demonstrations took place, with the teams, in both cases, located in different rooms. The first demonstration was conducted to test the platform and validate the injects before the real deployment in the Asclepios mission. In this demonstration, communication was conducted via Zoom, with cameras disabled and no dedicated communication loops. In the second demonstration, the pilot experiment, Asclepios’ COMMS software was used, enabling structured communication loops: one private loop between the FS, BME, and the astronauts; another between CAPCOM and the astronauts; and a third connecting the MCC roles.

This second test also incorporated delay constraints, including a bidirectional 3-second communication delay. Additional latency caused by network conditions and limited bandwidth frequently extended the delay in displaying vital signs data to more than 3 seconds. Additional factors included the physical separation between MCC and the astronaut crew, as well as the isolation of the astronaut crew. The pilot experiment was conducted on the 9^th^ day of astronauts isolation inside Sasso San Gotthard Fortress with 2106.0 meters of altitude and 6/7ºC.

### 2.4 Evaluation

This training platform was designed to assess and develop five key competencies: medical knowledge, procedural knowledge, communication, task load, and teamwork during a complex medical emergency.

The medical knowledge component covered three stages of medical emergency response: primary survey, secondary survey, and definitive care. Procedural knowledge encompassed crew roles and communication, as well as systems and procedural operations. Both competencies were evaluated through multiple-choice questions integrated into the simulation.

Communication performance was assessed both by the quality of decisions and by measuring response times, which were automatically recorded by the system. Each participant had five minutes to submit their decision via the platform. If the time limit expires, a one-point penalty is applied to the participant’s final score, followed by a one-minute grace period. Failure to respond within this additional minute triggers the system to automatically select the lowest-scoring option and advance to the next decision.

Task load was measured using NASA’s Task Load Index (NASA-TLX)[24], completed by each participant at the end of the simulation. Teamwork was evaluated separately for the astronaut crew, the MCC crew, and the combined team using three distinct Team Emergency Assessment Measure (TEAM)[25] surveys completed by the supervisors. Supervisors also qualitatively assessed situation awareness (SA).

## 3. Theory

### 3.2 Design and Development

#### 3.1.1 Management of a CVA in space

In an ischemic stroke—which accounts for nearly 90 % of cases—brain cells rapidly die from lack of oxygen and nutrients, whereas a hemorrhagic stroke damages tissue through increased pressure from leaked blood. Strokes are also classified by the location of the blockage or bleed. Prompt treatment is critical, as delays can lead to permanent disability or death; symptoms range from facial or limb weakness and numbness to sudden headache, vision loss, or speech difficulties[26].

Prolonged microgravity elevates intracranial pressure and redistributes cerebrospinal fluid, while hypovolemia-driven hemodynamic shifts remodel cerebral vessels—manifesting as 17–30% increased carotid stiffness and a 12% rise in intima-media thickness[19], [27]. Deep-space ionizing radiation also damages the vascular endothelium, increasing ischemic susceptibility [27]. Ground analogs, created with a 6° head-down tilt and mild hypercapnia, show significant reductions in cerebral blood flow, which raises the risk of transient ischemic events [19]. In-flight jugular venous stasis, with documented thrombosis, further heightens the risk of cerebral venous thrombosis. Combined with chronic hypercapnia, confinement stress, altered microbiomes, and high launch/landing G-loads, these factors synergistically impair endothelial function and foster prothrombotic states, creating a uniquely hazardous environment for ischemic stroke in deep space [19].

EIMO is supported by five core components [10]:

1. pre-mission planning
2. acute and emergent management decision-making
3. prolonged medical management decision-making
4. supplies and resource management
5. task load management

Two core EIMO components are (2) acute and emergent management decision-making and (5) task load management. The first involves ensuring onboard caregivers maintain peak readiness through structured simulation drills, ongoing knowledge assessments, just-in-time training, and integrated clinical decision-support tools—so they can stabilize patients until definitive ground guidance arrives. The second addresses the elevated cognitive demands of autonomous medical operations by deploying robust, intuitive decision-support systems that manage inventory control, medical records, proactive patient monitoring, diagnostic algorithms, treatment planning, and procedural reminders [10].

Preparedness for in-flight CVA emergencies includes pre- and during-flight astronaut training, development of emergency management protocols, and assembly of medical kits with appropriate equipment and supplies.

<u>Medical training</u> is essential for all crew members, flight surgeons, mission control support staff, and ground support personnel. Starting from their astronaut candidate year, crew members learn first aid, cardiopulmonary resuscitation (CPR), altitude and hypoxia drills, and medical equipment operation, tailored to mission specifics. This training includes the use of on-board medical kits, emergency interventions, and the study of space-specific physiology and toxicology [28].

CMOs receive specialized training in human physiology, behavioral health, diagnostics, pharmacology, and countermeasures [28]. On the International Space Station (ISS), CMOs complete hands-on training in med-kits, vital signs, catheterization, intravascular access, medication administration, and wound repair, all with ground support [28].

Pre-mission psychological training covers topics like isolation, group dynamics, cultural integration, family support, and behavioral health challenges for commanders, CMOs, crew members, ground personnel, and families. Physiological training, including carbon dioxide exposure and high-G adaptation, prepares them for environmental stressors and risk mitigation [29].

Flight surgeons assigned to support space programs must follow program-specific certification plans that include mission control operations, advanced cardiac life support (ACLS), advanced trauma life support (ATLS), aerospace physiology, hyperbaric medicine and emergency mishap response; parallel to this, all Medical Operations personnel in the MCC are trained and certified according to these tailored plans, while supervised training programs extend to flight directors, medical consultants, and other support staff who require proficiency in space medicine and flight medical procedures [28].

##### Emergency management protocols

Regarding stroke management in space, the current protocol is to stabilize the patient and start preparations for a return to Earth as soon as possible [19].

##### Medical kits in support of Cerebrovascular Stroke

To maintain airway patency and support ventilation in the event of neurologic deterioration, crews employ bag–valve–mask devices alongside oral. Circulatory stabilization is achieved with intravenous fluid administration sets—including a range of catheter gauges, infusion lines, and pressure infusers—paired with emergency medications such as antihypertensives to optimize cerebral perfusion. While the ISS pharmacy contained enoxaparin, no anticoagulation-reversal agent is available[30], [31].

A portable Electrocardiogram (ECG), pulse oximetry, and non-invasive blood pressure cuffs provide continuous monitoring of cardiac rhythm, oxygen saturation, and blood pressure. Instead of Computed Tomograph (CT) or Magnetic resonance imaging (MRI), point-of-care ultrasound serves as the primary diagnostic tool, using transcranial and cervical Doppler windows to assess vessel patency and infer intracranial pressure, thereby guiding treatment decisions in real time[31].

The medical kits used in this simulation are the ones currently in use in the ISS, 3.001 Medical KIT – Contents and Reference [32], — and other components retrieved from the ISS Crew Health Care Systems (CHeCS) Medical Hardware Catalog [31].

In the context of a medical emergency, the response phase begins at the moment the crisis unfolds: clinicians must rapidly diagnose the condition, initiate treatment, and actively manage the situation in real time[33].

On space missions, the response phase involves not only urgent clinical interventions but also the sustained physical and mental resilience of both astronauts and the MCC crew. Prolonged stress, fatigue, isolation, and residual injuries create a uniquely demanding environment that can impair decision-making and operational performance.

#### 3.1.2 Full-Scale Exercise

This type of exercise takes place in a realistic setting, utilizing resources to coordinate and respond to a planned event without disrupting infrastructure or risking public safety. It demands extensive resources for planning, execution, and evaluation compared to other exercise types [20].

Timed scenario “injects”, role-based consoles, and integrated audio–visual data feeds are used to test multi-agency coordination, command and control, and crisis decision-making under stress. Participants operate in real-time, managing information, executing response protocols, and collaborating across agencies, which helps identify coordination gaps and validates plans [14]. These exercises provide participants with the necessary resources and clear responsibilities, enabling them to assess situations and execute coordinated actions effectively and confidently[20].

#### 3.1.3 Coding of the platform

Web applications are software systems that run in a browser and are delivered over a network, eliminating the need for local installation. They are built with standard technologies such as JavaScript, and once loaded, execute locally, enabling platform independence and reducing deployment complexity [34].

Their architecture typically follows a client–server model: the client (browser) handles presentation and interaction, while the server manages business logic, data storage, and responses, with communication occurring via Hypertext Transfer Protocol (HTTP/HTTPS) [35]. This model streamlines development by centralizing updates and data management, while supporting scalability and cross-platform access— features aligned with best practices in modern web application design [21].

### 3.2 Demonstration

#### 3.2.1 Lunar Gateway Mission Conditions

The Orion vehicle will transport four-person crews —two women and two men— between Earth and the Gateway in a near-rectilinear halo orbit. Mission durations are planned to range from 10 days to up to 90 days, with each transit taking about four days [36]. Once in cislunar space, crews operate under a “deep-space lite” regime: despite a light-time delay of only 1.3 seconds, limited bandwidth and latency can stretch two-way communications to around 10 seconds, requiring a mix of synchronous and asynchronous exchanges.

Constraints on mass, volume, and power demand strict justification for all medical hardware and consumables. With no in-situ resource use or resupply during crewed phases, all medical equipment and pharmaceuticals must be pre-deployed, stored for the mission’s duration, and resistant to radiation, humidity, and elevated CO_2_ levels [36].

In the event of a medical emergency beyond onboard capacity, evacuation to Earth could take up to two weeks, obligating the medical system to sustain the crew during transit. The Orion vehicle, with no dedicated medical space, and the Gateway, with stowage and reconfigurable care areas, impose distinct constraints on volume, power, and environmental control [10], [36]. NASA Standard 3001 Revision 1A [37] defines five Levels of Care (I–V); Gateway missions are generally aligned with Level IV, which includes advanced life support, limited surgical and dental care, and imaging, though mission design may require deviations.

To meet these demands, at least two crew members are expected to serve as CMOs, though the precise level of training required remains uncertain [36]. Additional training is considered essential [10], given long evacuation times and the inability to resupply. The onboard medical system must support accurate diagnoses, anticipate the queries of ground specialists, and minimize communication loops. If a CMO becomes incapacitated, the system will require either a highly trained backup or autonomous capabilities to safeguard the crew[36].

#### 3.2.2 Realistic Simulations in Training

Analogue missions are Earth-based simulations of space missions conducted in isolated, confined, and extreme (ICE) environments, such as Antarctic stations and underwater habitats. They aim to validate technologies and human factors for long-duration exploration [38], [39]

Its participants use detailed “working cards” to complete multidisciplinary tasks, enhancing skills in teamwork, communication, decision-making, problem-solving, and time management despite limited resources and communication delays. These missions also serve as proving grounds for new hardware and operational concepts, contributing to the advancement of their Technology Readiness Level [40].

SRs function as command centers that integrate information and communication systems like disaster-management headquarters. Decision-makers use real-time monitoring to allocate resources effectively, minimizing damage and coordinating responses to emergencies [41].

### 3.3 Evaluation

#### 3.3.1 Medical and Procedural Knowledge

According to Robert et al. (2018) [33], medical emergency management “*consists of a rapid primary survey with simultaneous resuscitation of vital functions, a more detailed secondary survey, and the initiation of definitive care*”. This way, medical knowledge is divided into three categories: primary survey, secondary survey, and definitive care. The primary survey is based on the ABCDE framework of trauma assessment, which prioritizes [33]:

1. Airway protection with cervical spine stabilization
2. Breathing and ventilation
3. Circulation with haemorrhage control
4. Disability, referring to the rapid evaluation of neurological status
5. Exposure/Environment, to uncover hidden injuries while preventing hypothermia

This assessment is deliberately rapid and can often be completed in under 10 seconds. By asking a patient to state their name and describe what happened, if possible, or a person who found the patient, clinicians gain immediate insights. Conversely, an absent or abnormal response flags potential compromise in one or more of these domains and requires immediate intervention. The primary survey is not focused on defining specific injuries but rather on recognizing physiological threats to survival and addressing them in strict priority order [33].

Once resuscitative efforts are underway and vital functions show improvement, clinicians proceed to the secondary survey. This stage involves a systematic head-to-toe evaluation, including a comprehensive set of vital signs, detailed medical history, and a full physical exam. The goal is to identify injuries or conditions that may not have been immediately apparent but could deteriorate if untreated. Importantly, the secondary survey should never delay or interfere with ongoing life-saving interventions initiated during the primary survey[33].

The final stage of definitive care aims to stabilize and resolve a patient’s underlying conditions. In terrestrial trauma, this includes surgeries and intensive care. However, in spaceflight, definitive care is limited by available medical resources. NASA protocols focus on stabilization, symptomatic management, and preparation for evacuation, acknowledging that some conditions may exceed onboard treatment capabilities [33].

Procedural knowledge, which can be organized into two domains: (1) Systems and Procedural Expertise, covering familiarity with equipment, algorithms, protocols, and technical execution of interventions; and (2) Crew Roles and Communication, which emphasizes coordination, information flow, and the integration of decisions across the astronaut crew and mission control. In isolated and resource-constrained environments such as deep-space missions, these two domains are inseparable: technical competence without effective communication can lead to mismanagement of emergencies, while excellent teamwork cannot compensate for inadequate system knowledge[42], [43], [44].

#### 3.3.2 Task-load assessment

Zimmerman (2017) [45] refers to task-load as “*a measurement of human performance that broadly refers to the levels of difficulty an individual encounters when executing a task”*. The NASA-TLX is one of the most widely used instruments for evaluating subjective workload. Rather than focusing solely on objective task outcomes, it captures how demanding a task feels to the individual while performing it. The method assesses workload across six distinct dimensions, which together provide a composite measure of mental and physical strain [46], [47]:

1. Mental Demand – the degree of cognitive activity required, such as thinking, problem-solving, or decision-making.
2. Physical Demand – the level and intensity of physical effort needed to complete the task.
3. Temporal Demand – the perceived pressure of time constraints or deadlines during task execution.
4. Effort – the amount of energy the individual must invest to sustain performance.
5. Performance – the participant’s judgment of how successfully they accomplished the task.
6. Frustration – the extent of negative feelings such as stress, irritation, or discouragement, versus positive feelings of satisfaction or security.

After completing a task, participants provide ratings for each dimension using a 20-point interval scale, anchored from low (1) to high (20). These ratings are then combined to generate an overall workload index, reflecting the participant’s subjective experience of task difficulty [46], [47].

#### 3.3.3 Teamwork evaluation

While collaboration is fundamental to the success of any organization, it becomes vital in time-critical medical emergencies, where patient outcomes often depend on how well team members integrate their actions under pressure. In such contexts, the efficiency of communication, leadership, role allocation, and situational awareness directly determines whether life-saving interventions are delivered on time [25], [48].

To systematically evaluate these dynamics, the TEAM has been used to assess non-technical skills. This tool consists of 11 items rated on a 5-point Likert scale, covering three core domains[25]:

- Leadership (e.g., maintaining standards, directing and coordinating activities),
- Teamwork (e.g., communication, cooperation, adaptability, prioritization),
- Task Management (e.g., resource utilization, monitoring progress, maintaining a global perspective).
- Overall performance on a scale from 1 to 10.

#### 3.3.4 Situation Awareness qualitative assessments

Endsley (2021) [49] defines SA as an individual’s understanding of the current situation, a skill central to decision-making in high-risk domains such as aviation, emergency management, and health care. As a core non-technical skill, SA enables the perception of key cues, comprehension of their meaning, and anticipation of future states under pressure.

While quantitative SA measures exist, they risk disrupting performance by diverting cognitive focus. To maintain ecological validity, this study employed qualitative assessment through structured observation of both MCC and astronaut teams by two independent supervisors. This method captured behavioural indicators of SA, including information sharing, anticipation of problems, and responsiveness to evolving cues, within the natural flow of the simulation.

## 4 Results

### 4.1 Design and development

#### 4.1.1 Simulation scenario

The simulation was structured around two components: injects and decisions. Injects provide external inputs (such as updates or communications) accessible to all participants, reinforcing authenticity and situational awareness. Decisions required participants to actively engage with the scenario, either through multiple-choice questions that explored alternative predefined pathways or role-specific directives that reflected specialized responsibilities and authority in real-world operations.

Psychological strain was built into the design by layering stressors: complex events paired with strict time limits, fatigue induced by a two-hour high-intensity exercise, and isolation simulated by embedding astronauts already living under analogue lunar mission conditions.

The storyline began with Inject 1 and progressed through Decisions 1–15, with Decisions 12 and 15 serving as key branching points that defined six possible scenario pathways. Inject 2 followed; its timing is dependent on Decision 12, and in pathway 12(A), Decision 16 was added. Inject 3 appeared at 11:05 regardless of prior choices, with optional Decision 22 included when Decision 15 was not answered with option A. The simulation consistently concluded at 11:45, ensuring convergence across pathways.

The central medical emergency was an ischemic stroke during EVA on the Lunar Gateway. Midway, an additional stressor was introduced: a potential object collision with the station within one hour. Some pathways also produced a secondary complication, a minor pneumothorax.

Scenario design was informed by Earth-based guidelines, specifically the American Heart Association’s Stroke Algorithm [50] and *Tintinalli’s Emergency Medicine* [26]. The case simulated a middle cerebral artery infarction, the most common type of cerebrovascular accident, typically presenting with right hemiparesis, right central facial palsy, right hemisensory loss, and aphasia [51].

#### 4.1.2 Medical and Procedural Knowledge

The simulation advanced only to the stage of diagnosis and treatment selection, marking the start of definitive care. At each decision point, participants could also indicate insufficient information to support diagnosis or treatment.

The guidelines were adapted to the scenario, with Figure 3 mapping decision blocks to phases of emergency response. Due to external stressors, the phases overlapped and recurred.

**Figure 3.**
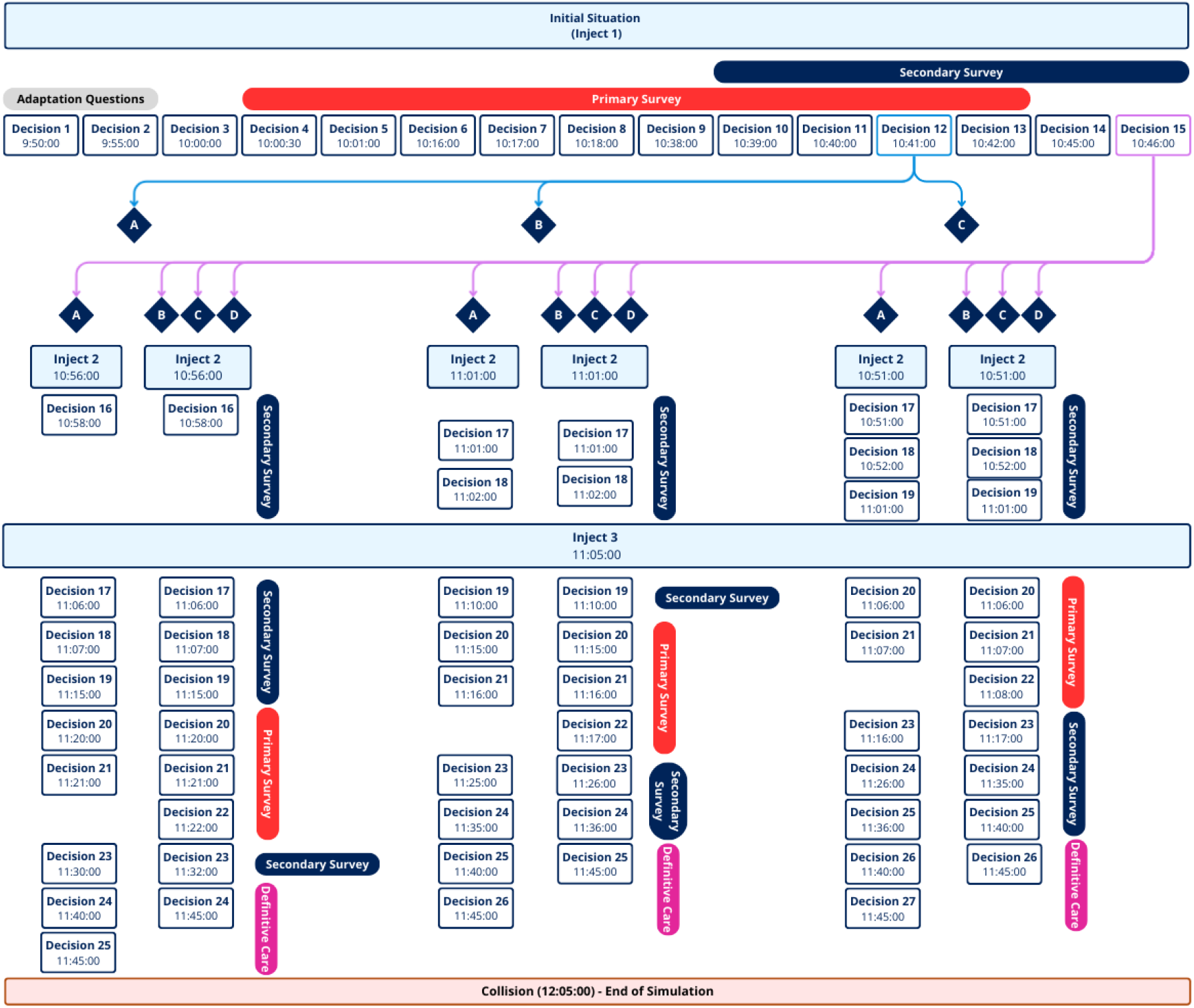
Scheme of the storyline blocks o decision divided in the medical emergency response phases

### The Simulation Story

The simulation begins with the first inject: “*On the 17*^*th*^ *mission day, an EVA is planned to add a component to a radiation data collection device. Preparations are complete, and two astronauts are outside performing the task*.” Day 17 was chosen to allow three days of travel and 15 days for adaptation, by which time astronauts are expected to have overcome space motion sickness and adjusted to microgravity. Unlike the ISS, the Gateway is outside Earth’s magnetic field and more exposed to radiation; hence, accurate measurements require placing the device outside the station.

The initial phase includes two adaptation questions, allowing participants to practice their responses and communication while the crew continues with routine tasks with the support of their supervisor. The medical emergency begins in Decision 3, when FE-1 reports experiencing numbness in their right arm. By Decision 6, symptoms are recognized as a potential sign of a stroke, prompting a primary survey. As the affected astronaut is outside, he and his partner must return inside, while the MCC begins a secondary survey, including neurologic tests and kit preparation. In Decision 9, facial asymmetry confirms suspicion. Decision 12 requires the Flight Director to choose a repressurization method, critical to stay within the 60-minute window. Standard repressurization takes around 15 minutes, but in an emergency, it can be reduced to 14 seconds at a rate of 1 psi/second [52]. In Decision 15, the FD must remind the EVA crew to breathe frequently, preventing pneumothorax if the patient becomes confused[53].

Inject 2, then inform participants that pressurization is complete. If Decision 15 was option A, EV1 presents confusion; otherwise, pneumothorax symptoms may occur, ranging from chest pain to shortness of breath. If Decision 12 was option A (10 minutes to 12 psi), Decision 16 adds that the hatch cannot open until pressure reaches 14 psi. From Decisions 16–19, the crew doffs EV1’s suit, records vitals, performs neurologic tests, and checks temperature. At 11:05, Inject 3 adds a collision alert: Probability of impact within one hour, with Pre-determined Debris Avoidance Maneuver (PDAM) unavailable. The crew must now manage both the medical emergency and prepare the Crew Return Vehicle (CRV) for isolation.

Between Decisions 20–22, the primary survey reappears as treatment relocates to a secure area. Decision 22 (conditional on Decision 15) addresses pneumothorax management. Medical participants continue the secondary survey—selecting kits for transfer, ordering glucose and electrolyte tests, and applying the NIH Stroke Scale (NIHSS) (score: 8)— while the non-medical group prepares the CRV. Without imaging, stroke mimics had to be excluded clinically, with assessments (e.g., Face drooping, Arm weakness, Speech difficulty, and Time to call for medical help (FAST), NIHSS) provided as decision support.

The scenario concludes with a final diagnostic decision for EV1, with “insufficient information” as an option. In Scenarios 12(C) & 15(A), participants also select a treatment option. The last inject emphasizes imminent collision, with 20 minutes remaining, requiring operational decisions. Finally, participants answer an opinion question: “*EV1 had an ischemic stroke, and the object collided with the station, causing a small undetected leak. Should the crew return to Earth, knowing the trip takes three days?*” Though unscored, responses assessed SA and reasoning. For more information, consult Appendix A.

#### 4.1.2 CHIRON platform

Users accessed the CHIRON platform via a login page, registering with a profile type code that identified them as participants or supervisors.

Figure 4 shows a simple flowchart of the platform. Upon login, the welcome page displayed project information, role identification, and three options: *New Simulation, Running Simulations*, and *Past Simulations*. Only supervisors could create simulations, assign participants to one of eight roles, and start the session once all roles were filled. Participants then entered the simulation, where each page showed the role, simulation name, and constant elements (decision number, event time, descriptive text, and a five-minute countdown). Responses were synchronized, with late or missing answers penalized.

**Figure 4.**
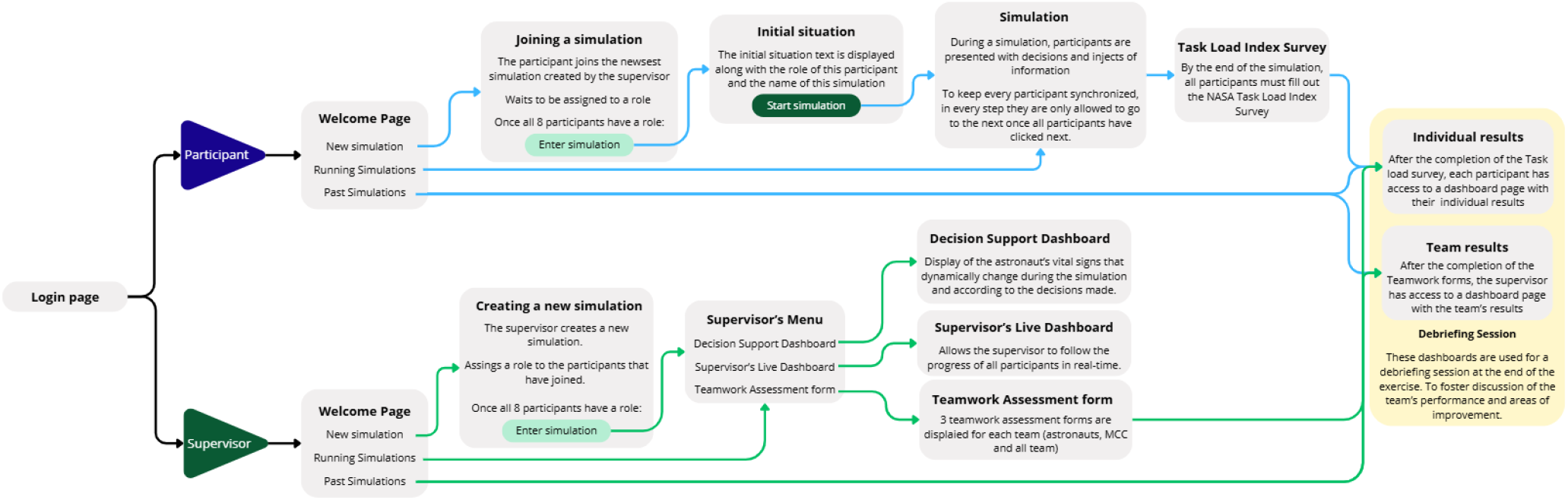
CHIRON Platform flowchart

**Figure 5.**
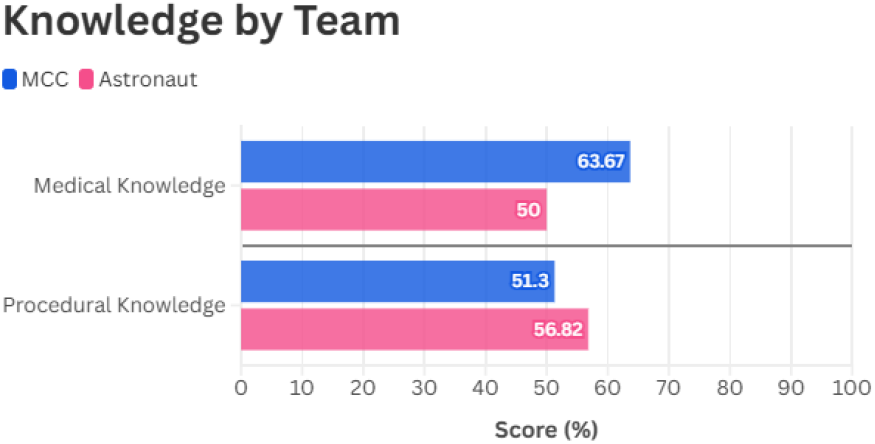
Percentage of Medical and Procedural knowledge by Team

**Figure 6.**
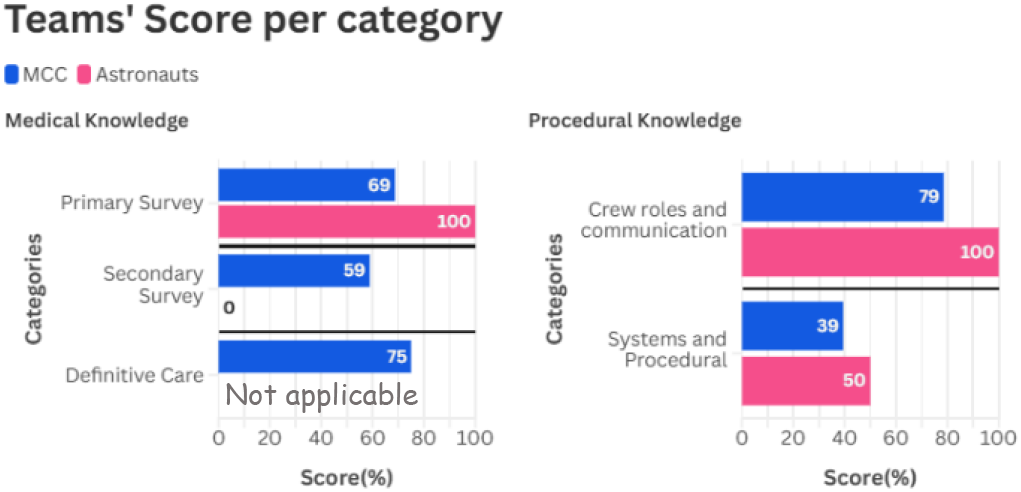
Percentage of the teams’ score per category of knowledge

The simulation was structured around injects (informational updates) and decisions (multiple-choice or role-specific directives). Supervisors had access to three tools: the vital signs dashboard (real-time astronaut vitals), the supervisor dashboard (real-time overview of all roles and statuses), and the teamwork assessment form.

At the end, participants completed the NASA-TLX survey and received an individual results dashboard with decisions, scores, graphs of medical/procedural knowledge, and task-load charts. Supervisors could also review team results, including aggregated scores, teamwork performance, and workload data. Both individual and team reports were exportable as PDFs. Results from completed simulations could be re-accessed at any time via the *Past Simulations* menu.

### 4.2 Demonstration

#### 4.2.1 Pre-pilot

The pre-pilot focused on testing CHIRON’s functionality and storyline feasibility rather than formal performance assessment. Participants had varied backgrounds; four had relevant expertise, including an FD, an intensive care physician as FS, and two medical students acting as Commander and FE-2. All received prior training in standardized communication protocols.

Conducted at the IDeaS Laboratory, NOVA School of Science and Technology, the pre-pilot used a longer storyline with 43 decision points, preventing full completion—the team reached only Inject 2 after two hours. Platform issues emerged, particularly delays caused by simultaneous answer submissions that overloaded the database.

Despite these constraints, participants engaged effectively with the scenario and applied appropriate emergency procedures. Notably, they performed emergency repressurization at 1 psi/second and instructed EVA astronauts on safe breathing techniques, ensuring re-entry of the injured astronaut without pneumothorax.

#### 4.2.2 Pilot experiment

The pilot experiment served as the formal testing of the platform within an analogue lunar mission environment replicating the operational constraints of lunar exploration. Eight participants were selected: four astronauts and four MCC members, all students ranging from bachelor’s to PhD and trained in communication and operational procedures. The MCC team included a biology PhD student as FD, a bachelor’s student as CAPCOM, a medical student as FS, and a master’s student in human factors as BME. The astronaut crew consisted of a medical student as CMO, IV2, a palaeomimicry PhD student as Flight Engineer-2 (IV1), and two aerospace engineering students as Flight Engineer-1 (EV1) and Flight Engineer-3 (EV2).

The experiment lasted around 3 hours, including a project briefing, a guided walkthrough of the platform, the simulation itself, and a short debriefing. The simulation phase lasted 2 hours and 28 minutes. and followed Scenario 12(C) & 15(A).

### 4.3 Evaluation

#### 4.3.1 Pre-pilot

The pre-pilot enabled an initial assessment of CHIRON’s functionality and usability, informing improvements in later versions. Participants recommended automatic page refresh, supervisor-only role assignment, optimized database requests, and a “Past Simulations” option for accessing previous results.

Storyline feedback highlighted that 43 decision points were too many for the three-hour format, leading to a reduced 27-point version. Participants also noted insufficient role-specific information, which was later addressed by adding tailored directives to increase realism and engagement.

Supervisors observed that efficient communication and clear leadership improved coordination and decision quality, while task overload increased stress and fatigue. Participants without prior training exhibited more hesitation and sometimes overlooked details, such as fully reading prompts, which negatively affected performance.

#### 4.3.2 Pilot experiment

Unlike the pre-pilot, the pilot formally evaluated knowledge, workload, and teamwork. The simulation was completed, following Scenario 12(C) & 15(A).

A limitation occurred at Decision 12 when a 10-minute outage, probably caused by internet failure, disrupted communication between the frontend and backend. The platform needed a manual reboot by the supervisor, which interrupted the decision-making process and could have impacted situational awareness and engagement.

### Medical and Procedural Knowledge

Regarding the two types of knowledge assessed (medical and procedural), neither the MCC, astronauts, nor the All team obtained a score above 70%. The MCC achieved the highest score in medical knowledge (63.67%), whereas the astronaut team obtained the lowest score in this category (55.56%). For procedural knowledge, the astronaut team had the highest score (56.82%), followed by the MCC (51.3%).

In the medical knowledge field, although astronauts scored 100% in the primary survey category, they were unable to obtain any points (0%) in the secondary survey category, which comprised the remainder of their medical knowledge. For the MCC team, the Secondary Survey was the lowest-scoring category (59%), while the highest score was obtained in the Definitive Care (75%), and finally, the Primary Survey scored 69%.

In Crew Roles and Communication, MCC scored 79% and astronauts achieved 100%. In contrast, in Systems and Procedural Expertise, both teams achieved results not higher than 50%, with MCC at 39% and astronauts at 50%.

Figures 7 and 8 present the category-level scores for each role. In procedural knowledge, the BME, FS, FD, FE-1 (EV1), FE-2 (IV1), and the Commander all achieved 100% in the Crew Roles and Communication category. The only exception was CAPCOM, which obtained 25% in this category. In Systems and Procedural Knowledge, the highest score was recorded by FE-2 (IV1) (100%), while the BME and FE-1 (EV1) scored 0%, representing the lowest performance.

**Figure 7.**
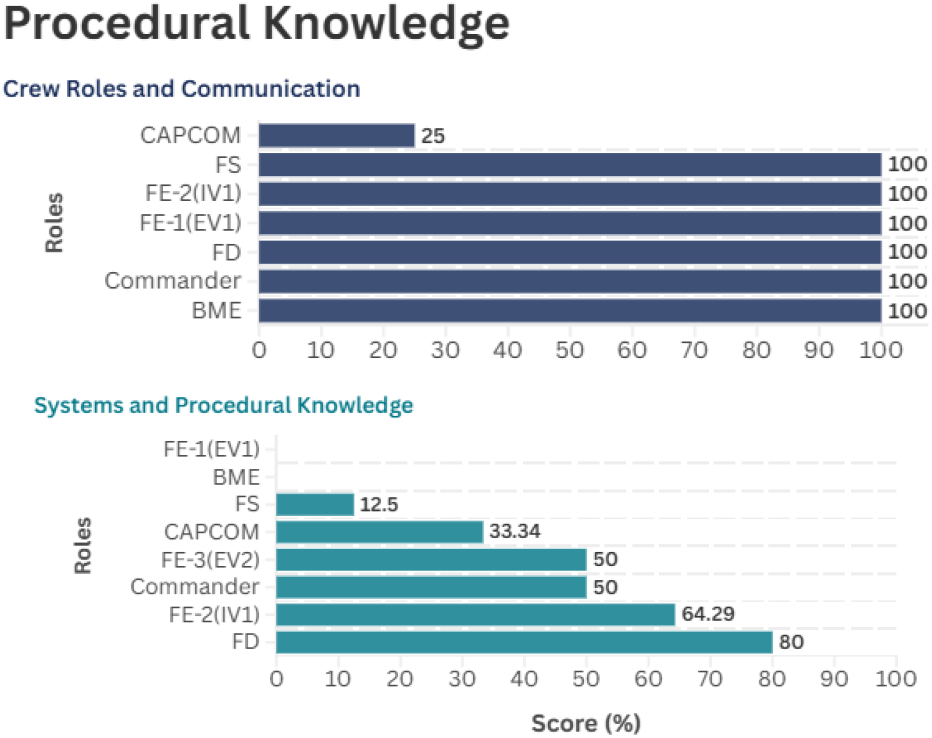
Percentage of Procedural Knowledge categories per role

**Figure 8.**
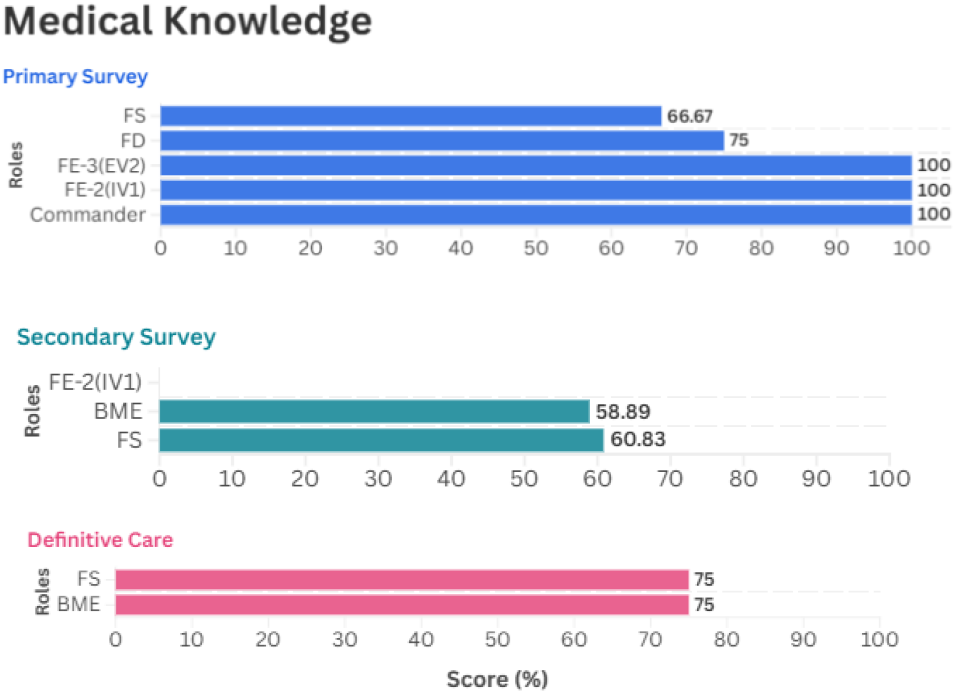
Percentage of Medical Knowledge categories per role

In medical knowledge, the BME and FS achieved comparable scores across most categories, except for the Primary Survey, where the BME had no related decision. The Primary Survey was also the only medical category in which four additional roles contributed: the FD achieved 75%, while FE-2 (IV1), FE-3(EV2), and Commander reached 100%.

### Answer time

In the simulation, the answer time for each decision was recorded. Figure 9 presents the average answer time per decision for MCC, the astronaut team, and the combined team. MCC recorded a higher average answer time than the astronaut team in 44% of the decisions, while the astronaut team recorded higher values in 40% of the decisions. In 16% of the decisions, both teams had similar average answer times. MCC registered the highest average answer times overall, with 347.50 seconds in Decision 7 and 292 seconds in Decision 15. Across the simulation, both teams showed peaks in answer time at completely different decision points, particularly in Decisions 3, 7, and 20. The key decisions, 12 and 15, were answered only by the FD, resulting in an average answer time of 0 for the astronaut team. In some decisions, certain roles only received instructions without having to submit a decision, which influenced the average answer time for those decision points.

**Figure 9.**
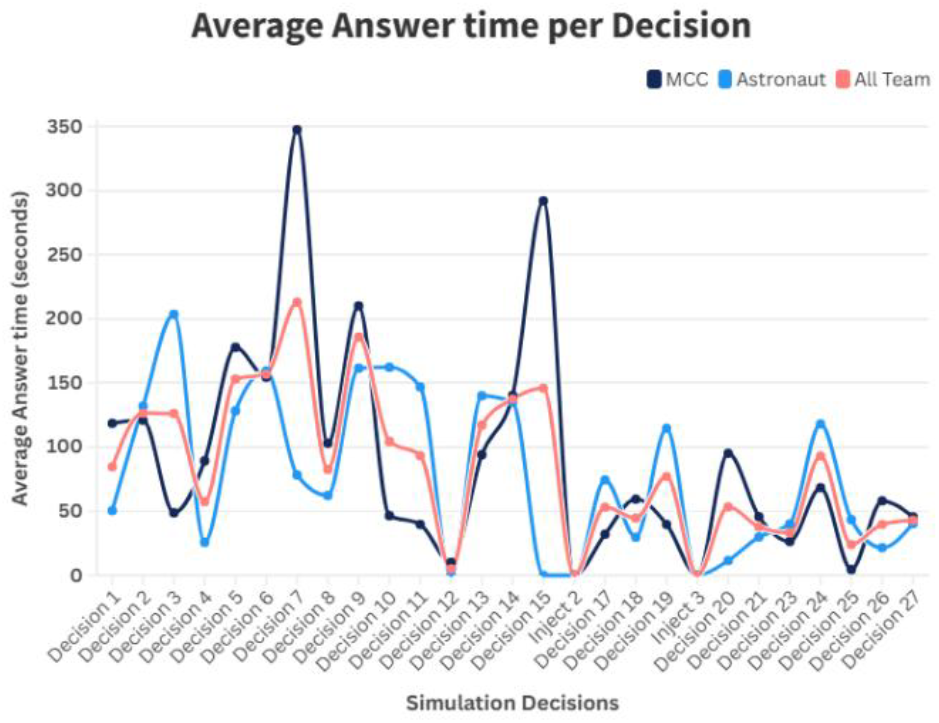
Average answer time per decision

### Task-load

Immediately after the simulation ended, all participants completed the NASA-TXL survey. Figure 10 shows the task-load average answers given by each team. Both teams considered the task low in physical task-load but higher than 5 points in all other domains. The astronaut team considered the mental and frustration load of the task particularly high. In comparison, the MCC team considered the effort and temporal load to be notably higher. An interesting fact is that MCC gave much higher points to the temporal load (18.75) than the astronaut team (10.5).

**Figure 10.**
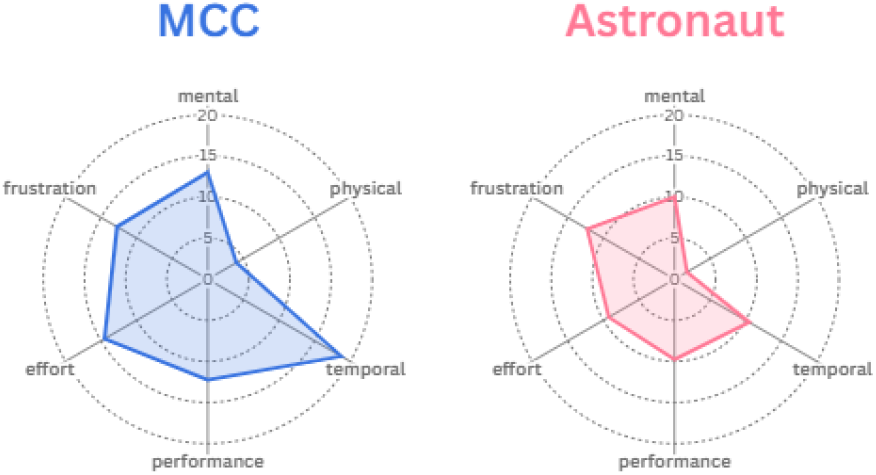
Taskload per team

**Figure 11.**
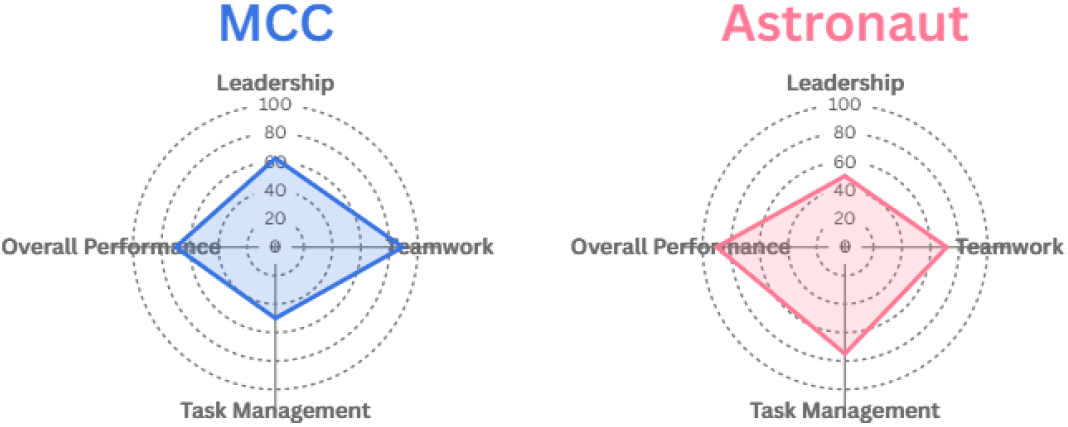
Percentage of Teamwork score per team

### Teamwork

Two supervisors assessed teamwork during the simulation. Leadership scores were low in both teams, with MCC at 62.5% and the astronaut team at 50%. Task management was 50% for MCC and higher for the astronauts at 75%. Teamwork was the highest-rated domain, with MCC scoring 89.3%. Overall performance was rated as good, with MCC at 70% and the astronauts at 90%.

## 5. Discussion

The simulations showed uneven performance: astronauts excelled in procedural tasks, MCC in medical knowledge, but overall scores remained below 70%. Communication was generally effective, yet leadership and task management were weak. NASA-TLX highlighted role-specific stress, with MCC under temporal pressure and astronauts reporting frustration.

### 5.1 Knowledge performance

Results showed heterogeneous performance, with neither MCC nor astronauts achieving an overall score exceeding 70%. Astronauts scored higher in procedural knowledge and MCC in medical knowledge: differences that reflected their academic backgrounds and limited training. Interpretation is further affected by the unequal number of decisions per role.

#### 5.1.1 Medical knowledge

The scenario began with an astronaut, FE-1 (EV1), reporting numbness in the right arm (Decision 3), leading CAPCOM to consult the Flight Surgeon (FS). This symptom could have been from strenuous EVA maneuvers or an underlying condition. Since transient limb numbness can occur during EVA due to mobility restrictions [54], this should have been ruled out before escalating to emergency procedures. However, the FS prematurely advised an EVA abortion. A possible explanation is the framing effect from the pre-simulation briefing, which could have influenced participants to view the initial symptom as pathological rather than operational, highlighting the impact of context on diagnosis.

### Primary Survey

Between Decisions 4 and 13, the team conducted the primary survey, during which the FS assessed the ABCDs through private communication with EV1. In Decision 6, the team recognized that the symptoms were unlikely to be task-related and instead indicated a possible medical emergency: the numbness did not resolve with rest, and weakness appeared in the same limb. This phase emphasized scene safety as a first step in basic life support. However, in Decision 7, the FD instructed EV1 to return alone to the crew lock, contradicting patient safety protocols that consistently emphasize that patients experiencing acute medical events, particularly neurologic ones, should never be relocated without assistance [55]. Given the distance between the two EVA astronauts and the 20-minute journey, keeping the patient in place until help arrived would have been the safer option.

The simulation design limited branching decisions, so not all choices impacted the flow of the scenario. Although EV1 received unsafe instructions to move alone, the system directed EV2 to assist and perform prehospital neurologic tests. At this point, the FS appropriately instructed EV2 to perform prehospital neurologic tests (e.g., FAST) to support a differential diagnosis. However, as highlighted by Rosenberg et al. (2025) [19], standardized tools like FAST or NIHSS for emergency neurologic assessment have not yet been adapted for use in a microgravity environment. Consequently, in this simulation, the outcomes of these algorithms were provided directly to participants rather than requiring them to perform the assessments, reflecting both the current limitations of space-adapted protocols and the need for future research to operationalize these tools in these environments.

In Decision 9, the observation of right-sided facial drooping indicated a cerebrovascular event, leading the FD to correctly abort the EVA and return both EV astronauts, prioritizing safety. In Decision 10, FE-3 (EV2) accurately answered regarding patient transport by staying close, monitoring EV1’s condition, providing reassurance, and updating CAPCOM.

By Decision 11, limitations in medical knowledge and situational awareness became evident when the FS suggested relocating the patient inside the station rather than treating near the crew lock, where the Crew Medical Restraint System (CMRS) could be fixed. This choice overlooked the well-documented risks of transporting unstable patients, as clinical studies show that movement can precipitate deterioration, particularly in neurologic or critically ill cases [55]. A safer approach and the correct answer would have IV astronauts bring the necessary medical kits closer, ensuring both reduced transport-related risk and a timelier medical response by treating them directly near the crew lock.

In Decision 13, the final step of the primary survey, the FS correctly identified the last known normal and acknowledged that treatment should ideally be initiated within the 60-minute therapeutic window to improve stroke outcomes[26]. Although there were no issues or specific decisions related to oxygen saturation, this is a critical parameter in stroke algorithms, but the MCC medical team failed to verify this information on the vital signs’ dashboard, reflecting a significant gap in situational awareness and protocol adherence. This omission deviates from the standard stroke protocols[50], which emphasizes monitoring and maintaining adequate oxygenation as a fundamental step in emergency stroke care[9].

Later in the scenario, at 11:05, Inject 3 introduced a second stressor: a potential collision with the Lunar Gateway within one hour. This shifted attention to both operational and medical decision-making. In Decision 20, the FD was asked how to respond to the collision threat, while the FS had to reconsider whether the treatment site was still appropriate. The FD correctly commanded isolation in the CRV as a contingency against depressurization. The FS likewise decided to relocate medical care to the CRV. Although both decisions were technically correct, the lack of team discussion regarding the risks and trade-offs associated with relocation limited collaborative reasoning and decision-making. Ideally, the FS should have explicitly justified the choice, and the FD should have probed further before confirmation.

This discussion reflects the trends in Figure 8. Notably, although with a higher number of questions for this category, the FS—despite being the most medically trained member of the team—scored lower than both the FD and FE-3 (EV2) during this phase. This finding highlights deficits in both the application of medical knowledge and situational awareness, raising concerns about overreliance on formal training without adequate contextual decision-making.

Together, these findings highlight the vulnerability of medical decision-making in spaceflight contexts, where role expectations may not align with actual performance under pressure. Reinforcing the importance of simulation-based training in a situation room context to strengthen situational awareness, ensure adherence to critical algorithms, and minimize risks associated with unsafe relocation of unstable patients.

### Secondary Survey

Since the simulation doesn’t reproduce an ideal scenario for medical emergency response, the secondary phase overlapped with the primary survey. This way, this phase progresses from decision 10 to 15 before Inject 2, overlapping with the primary survey. There were questions that the MCC medical team could perform in the background, such as developing a differential diagnosis list to help understand what other tests should be done to discard other possible diagnoses. During the movement of the EV astronauts inside the base, subsequent preparations involved selecting the appropriate medical kits. Here, at decision 14, FE-2(IV1), FS, and BME had difficulty interpreting the question, which specifically requested the identification of individual pieces of equipment, but the options given were medical kits. This made it difficult to interpret the question. However, this again highlights the limited knowledge on medical kits contents.

The answers to the two key decisions led to Inject 2, achieving the best possible repressurization time without secondary complications for FE-1 (EV1). At this point, however, this astronaut’s condition deteriorated further: he presented with confusion and was no longer able to participate in discussions. FS and BME appropriately prioritized the doffing of the EV1 suit, reflecting an understanding of the need to assist the affected astronaut rapidly. Nevertheless, subsequent actions revealed gaps in medical reasoning. In Decision 19, instead of restraining the patient with the CMRS, a vital measure that facilitates emergency medical maneuvers such as CPR in aerospace environments[5], they merely recorded basic vital signs without performing a neurological assessment (NIHSS) or measuring temperature, both of which are essential for understanding the complexity of the situation.

The omission is significant: managing acute neurologic emergencies, such as suspected stroke, requires not only physiological stabilization but also rapid exclusion of alternative diagnoses, so-called “stroke mimics,” including conditions such as meningitis, hypoglycemia, or seizure [26]. Omitting a basic work-up (including temperature, neurological exam, and relevant point-of-care testing) obscures diagnostic clarity and delays appropriate intervention.

In Decision 18, both the BME and FS responded, “*Wait to see the vital signs data*”, despite this information already being available on the dashboard. This response foreshadowed what became evident again in Decision 21, a persistent weakness in situational awareness. Both participants claimed insufficient vital-sign information to assess EV1’s physiological status, even though the dashboard clearly displayed key parameters, including heart rate, respiratory rate, blood pressure, Oxygen Saturation, temperature, NIHSS score, and electrolytes. The failure to integrate this information led to an unnecessary delay: the doffing of the EV2 suit was suspended until stability could be “confirmed,” despite dashboard data indicating that EV1 was already stable. By contrast, subsequent responses, such as ordering glucose and electrolyte testing and performing NIHSS assessment, were correct, and the team improved when re-asked to identify medical kits for treatment in the CRV. This pattern suggests that learning and adaptation occurred during the simulation yet also highlights the need for stronger training in information monitoring and integration to avoid overlooking readily available clinical data.

These shortcomings are reflected in the scores presented in Figure 8, where performance in the secondary survey phase remained low, with no role achieving more than 70%. Given that the secondary survey is a critical step in structured emergency response, these results highlight the risk posed by insufficient situational awareness, incomplete use of available tools (e.g., dashboards), and limited application of diagnostic algorithms. Together, these results emphasize the need for targeted training interventions to reinforce structured secondary survey practices, ensure proper use of medical systems, and enhance the integration of available clinical data into real-time decision-making.

### Definitive Care

The final decisions for the FS and BME highlighted inconsistencies in stroke care. Participants expressed a lack of information for a diagnosis, yet still chose to administer aspirin, assuming ischemic stroke was most likely due to its prevalence. This raises concerns because aspirin could be harmful in the case of a hemorrhagic stroke.

This ambiguity highlights two critical issues: first, the need for clearer diagnostic decision-making frameworks in space medicine training, particularly when advanced imaging is unavailable; and second, the vulnerability of medical decision-making under uncertainty and operational stress. Rosenberg et al. (2025)[19] emphasize, future long-duration space missions will require detailed stroke emergency protocols that go beyond simple stabilization and evacuation to Earth, providing structured guidance for diagnosis and management in-flight. This underscores the importance of training both astronauts and MCC staff in stroke algorithms and managing diagnostic uncertainty, including guidelines for withholding treatment.

#### 5.1.3 Procedural knowledge

This type of knowledge accompanied the development of medical emergency and the external factors that influenced the unwind of the story.

### Crew Roles and Communication

All roles except for FE-3 (EV2) contributed to decisions within this category, and nearly all—except CAPCOM (25%)—achieved the maximum score (100%). Interestingly, CAPCOM, the role responsible for coordinating communication between MCC and the astronaut crew, obtained the lowest score overall. While this may partly be explained by the greater number of decisions allocated to CAPCOM within this dimension, it also reflects gaps in knowledge of communication procedures and crew roles. Supervisor observations, however, offered a more nuanced picture: although CAPCOM’s transmissions were at times overly slow and detailed for the urgency of the emergency scenario, overall communication performance was assessed as satisfactory.

At first glance, these results might suggest effective communication across the team; however, closer inspection of answer times and interaction patterns reveals underlying weaknesses. Specifically, the MCC team demonstrated limited familiarity with the communication hierarchy that should be followed during medical emergencies. Instead of prioritizing input from the FS and BME, the members with medical expertise, greater emphasis was consistently placed on CAPCOM and FD directives. This dynamic indicates that decision-making was driven more by operational authority and ingrained communication habits than by clinical logic. Furthermore, participants’ behavior suggested that their primary goal was to submit timely responses within the simulation’s constraints, rather than fully engaging in collaborative reasoning to optimize medical management.

This raises important concerns for real mission contexts, where the pressure to meet procedural timelines may inadvertently overshadow the need for deliberate medical decision-making. These findings reinforce the necessity of embedded explicit training on communication hierarchies and role prioritization within space medicine protocols. Such training should ensure that medical expertise is not subordinated to operational authority in emergencies, and that decision-making reflects a balance between procedural efficiency and clinical safety.

### Systems and procedural knowledge

The scores presented in Figure 7 indicate generally low levels of systems and procedural knowledge across the team, with the notable exception of the FD, who achieved 80% and demonstrated a comparatively strong understanding in this domain. This was reflected in his ability to make the correct decision regarding repressurization—opting for emergency repressurization at a rate of 1 psi per second— supported by the rest of the team. This decision was critical, as it led to Inject 2 with the optimal repressurization time achieved. Subsequent procedural steps, such as hatch opening after repressurization, were also performed correctly.

The introduction of Inject 3 at 11:05, however, exposed more systemic weaknesses. When confronted with the added stressor of a potential collision with the Lunar Gateway, none of the participants could identify the CRV, requiring direct supervisor intervention to clarify its function. Although the FD subsequently issued the correct order to isolate in the CRV and the team successfully completed procedural preparations, this knowledge gap highlights a critical vulnerability: even fundamental spacecraft systems were not universally understood among participants.

Further operational decision-making revealed additional misconceptions. For example, participants suggested undocking the CRV before the collision event occurred, disregarding the need to confirm the impact first and to evaluate return windows to Earth.

In addition, they mis-sequenced several steps required in the final 20 minutes before the scheduled collision, reflecting procedural gaps in mission operations knowledge. These errors suggest that while baseline technical proficiency was sufficient for discrete, well-defined tasks, the team struggled when confronted with more complex operational contexts that required integrating medical and spacecraft system knowledge under time pressure.

Taken together, these findings highlight the uneven distribution of systems and procedural knowledge within the team and raise concerns about overreliance on a single role (the FD) to ensure correct system-based decision-making. This not only increases workload and cognitive pressure on one individual but also risks catastrophic failure if that role becomes impaired. For future missions, this underscores the importance of cross-training astronauts and MCC personnel in spacecraft systems, ensuring redundancy in critical knowledge areas and reducing dependence on single-point expertise.

### 5.2 Answer time

Figure 12 illustrates the answer times per decision for MCC crew members, organized by medical emergency response phase. Decisions 4 to 13 corresponded primarily to the primary survey but also incorporated elements of the secondary survey. Completing this block required almost 40 minutes, whereas the *Adult Suspected Stroke Algorithm* recommends that the primary survey be completed within 10 minutes [50]. This discrepancy highlights a significant performance gap compared to clinical best practices.

**Figure 12.**
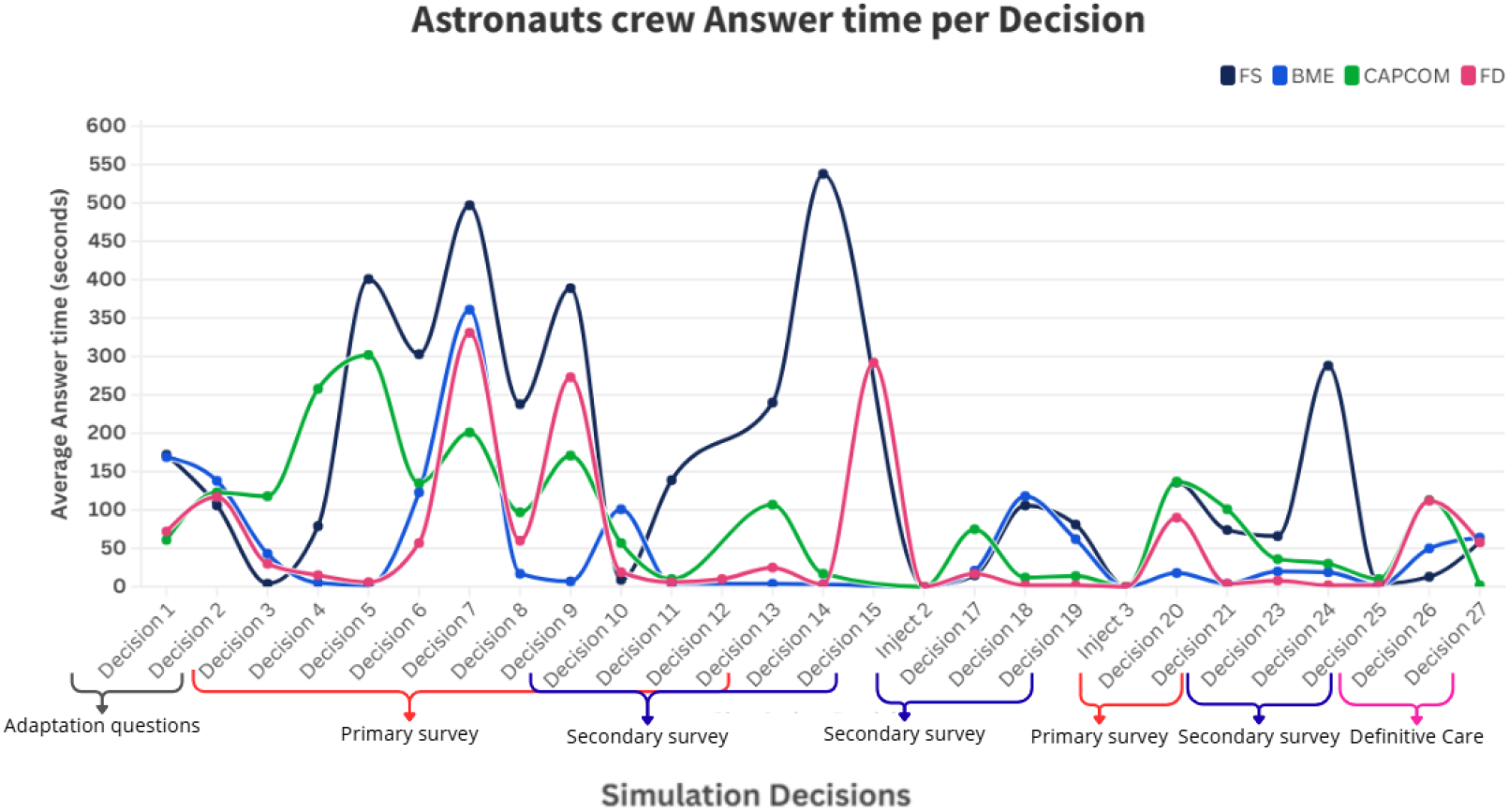
MCC crew members answer time per decision

Nonetheless, it is important to acknowledge that this was not an “ideal” emergency response environment: the crew was simultaneously tasked with aborting the EVA and managing re-entry, increasing cognitive workload, and diverting attention away from medical priorities. In reality, emergencies rarely unfold under optimal conditions, and this reinforces a critical point: preparation and training should not be designed for idealized scenarios, but rather for worst-case conditions where competing operational and medical demands converge.

Communication dynamics further contributed to delays. In Decision 3, a microphone malfunction caused a delay that the team passively endured instead of adopting alternative communication strategies. This incident reflects both the vulnerability of technical systems and a lack of adaptive problem-solving under operational stress—factors that, in a real mission, could exacerbate clinical risk.

Supervisor observations and response-time data suggest that the MCC prioritized directives from CAPCOM and FD over those from FS and BME. While this mirrors operational authority structures, it highlights insufficient awareness of medical communication hierarchies, where clinical expertise should be prioritized in emergencies. This is reflected in Figure 12, where FS response times were consistently higher than other roles, suggesting delays in integrating medical input into the decision-making process. Instead of engaging collaboratively to optimize patient outcomes, participants appeared primarily focused on responding within the simulation’s time constraints. This tendency raises concerns that, under real mission conditions, time pressure and procedural compliance may overshadow adaptive clinical reasoning and coordinated decision-making, with potential consequences for patient safety.

The secondary survey phase provides additional insight. Ideally completed within 25 minutes, this phase required nearly 40 minutes. Figure 12 identifies three decisions—14, 15, and 24—as particularly time-consuming. At Decision 14, FE-2 (IV1), FS, and BME struggled to interpret a question that asked them to identify individual pieces of equipment, although the available options were medical kits. Despite having been provided with the kit list one week prior, participants demonstrated limited knowledge of kit contents, resulting in confusion and delays beyond the maximum time allowance. In key Decision 15, similar challenges were evident: the team hesitated over which actions were most critical for the astronauts during repressurization, requiring 292 seconds to respond. Although they ultimately selected the correct answer, supervisor debriefs indicated that the decision was reached through a process of elimination rather than an understanding of the underlying physiological rationale. This gap suggests a limited understanding of pressure-related physiology, particularly in the context of hyperbaric training. It highlights a reliance on superficial reasoning rather than confident, evidence-based decision-making. Finally, Decision 24 once again involved the selection of medical kits, an inherently complex task that, despite prior exposure, remained a source of delay.

Taking together, these findings reveal how limited systems knowledge, communication inefficiencies, and high cognitive load interact to prolong decision-making across both primary and secondary survey phases. They underscore the importance of targeted training not only in medical algorithms but also in equipment familiarity, hyperbaric physiology, and communication hierarchies under stress. Addressing these vulnerabilities through simulation-based education and cross-training may be crucial to ensuring that astronaut crews and MCC teams can provide timely and accurate responses in real mission emergencies.

### 5.3 Workload and stress factors

NASA-TLX results indicated that both crews perceived the task as highly mentally demanding, with MCC members reporting greater temporal load while astronauts reported higher frustration. These perceptions are consistent with the scenario’s deliberate design, which layered multiple stressors, including strict time pressure, communication latency, and the simultaneous management of operational and medical emergencies. Such conditions are known to elevate cognitive workload and degrade performance in high-reliability teams, as shown in both aerospace and emergency medicine simulations[56], [57].

The divergence between MCC and astronaut workload profiles is particularly noteworthy. For MCC, the dominant stressor was time pressure, reflecting their responsibility for integrating multiple streams of information and issuing timely directives. For astronauts, frustration likely arose from technological limitations—such as communication failures and equipment interpretation—and from restricted autonomy, which can magnify stress in isolated medical emergencies. This asymmetry underscores how workload is shaped not only by role but also by context, even within the same mission scenario. More importantly, it highlights a critical implication for future deep-space missions: constraints on communication will demand greater autonomy from astronaut crews, reducing reliance on MCC directives and reinforcing the need for advanced training and decision-support tools to sustain medical independence.

These findings suggest two important implications. First, role-specific workload mitigation strategies should be incorporated into future training. For MCC, this may involve structured communication protocols and decision support tools to buffer against temporal load. For astronauts, emphasis should be placed on practicing adaptive problem-solving under uncertain conditions, thereby fostering resilience when equipment or communication systems fail. Second, the results suggest the potential value of incorporating stress inoculation training and workload monitoring into analog missions to quantify better and mitigate the impact of operational stress on medical decision-making.

Ultimately, while elevated workload and stress are unavoidable in real missions, targeted training interventions can reduce their negative impact. Simulation studies consistently demonstrate that crews who practice under high-stress conditions develop greater resilience and more effective teamwork, improving both task performance and patient safety outcomes [58]. Integrating these approaches into spaceflight medical training could therefore strengthen both MCC and astronaut readiness for emergencies beyond Earth orbit.

### 5.4 Teamwork and communication

Teamwork assessments identified cooperation as a relative strength but exposed persistent weaknesses in leadership and task management, particularly within the MCC team. These findings are consistent with evidence from other high-reliability domains, where the absence of a clearly designated leader and inconsistent adherence to communication protocols can undermine coordination, delay decision-making, and reduce overall team effectiveness[58]. In this simulation, hesitations in leadership structure appeared to reinforce role ambiguity, with participants prioritizing procedural completion over collaborative problem-solving.

A notable improvement was observed in the structured communication loops introduced in the pilot. Compared to the pre-pilot, which relied on ad hoc Zoom-based exchanges, the adoption of standardized communication channels markedly enhanced clarity and reduced message loss. By replicating realistic communication pathways, the pilot fostered better alignment between astronauts and MCC, supporting more efficient information flow even under time pressure.

Taken together, these results highlight that while baseline cooperation was strong, effective crisis response requires more than goodwill—it demands explicit leadership, adherence to structured communication protocols, and training in task management under stress. Strengthening these dimensions should therefore be a priority for future analog missions, both to increase ecological validity and to prepare crews for the leadership and communication challenges inherent in spaceflight medical emergencies.

### 5.5 Implications for future research

The findings underscore the CHIRON platform’s potential as both a research and training environment for deep-space medical operations. As lunar missions expand, medical autonomy will be central to mission resilience. Simulation-based systems such as CHIRON can (i) expose critical training gaps in medical and procedural domains, (ii) assess workload and stress management under realistic constraints, and (iii) provide structured opportunities to strengthen teamwork, communication, and decision-making. Coupling these platforms with physiological monitoring tools (e.g., EEG for early detection of neurological events) could further bridge the gap between analogue training and in-flight medical autonomy.

The study also revealed current limitations in space-adapted medical protocols, particularly for CVAs. Existing diagnostic algorithms are not operationalized for space environments, and the prevailing directive of “stabilize and return to Earth” is insufficient for ischemic stroke. Given that evacuation from lunar orbit can take up to two weeks, survival without severe neurological sequelae is unlikely in the absence of targeted intervention. Future research must therefore prioritize the adaptation of stroke protocols to spaceflight conditions, moving beyond stabilization toward sustained in-flight treatment strategies.

## 6. Conclusion

This study demonstrated the feasibility and potential of the CHIRON simulation platform for training both astronauts and MCC personnel in managing acute medical emergencies under deep-space mission conditions. The results highlighted not only essential strengths, such as participants’ ability to adapt during the scenario and the benefits of structured communication loops, but also significant limitations in medical knowledge, procedural execution, and situational awareness. These findings underscore that, even under controlled analogue conditions, operational stressors and communication hierarchies can undermine medical decision-making, emphasizing the need for targeted training in both technical and non-technical skills.

Several limitations must be acknowledged. The participant sample was composed exclusively of students with limited exposure to aerospace medicine and operational systems, limiting generalizability to professional astronaut or MCC populations. Furthermore, the absence of advanced diagnostic tools (e.g., neuroimaging) in the simulation restricted diagnostic fidelity, though this constraint realistically reflects the limitations of spaceflight medicine.

Future research should address these limitations by involving more diverse participant groups, including professionals with operational or clinical experience, and by incorporating objective stress biomarkers (e.g., heart rate variability, salivary cortisol) to link workload with performance outcomes. Improving the realism of the simulation through adaptive AI-driven branching storylines could also better capture the dynamic and uncertain nature of real spaceflight emergencies. This platform can also help determine the medical knowledge and equipment needed for specific space missions. Beyond astronaut training, the CHIRON system offers wider application for Earth-based disaster medicine, especially in settings where teams must manage limited resources, delayed evacuations, and high cognitive workload.

Ultimately, the results reinforce the importance of developing detailed, evidence-based medical protocols that extend beyond stabilization and evacuation, particularly for time-critical conditions such as stroke. Preparing astronauts and MCC teams for medical autonomy in deep space will require not only robust technical protocols but also systematic investment in simulation-based training, where communication, leadership, and decision-making under uncertainty are as rigorously assessed as clinical skills.

## Data Availability

All data produced in the present work are contained in the manuscript.

## Acronyms/Abbreviations

ISS: International Space Station
NASA: National Aeronautics and Space Administration
LEO: Low Earth Orbit
EIMO: Earth-Independent Medical Operations
CVA: Cerebrovascular Accident
FEMA: Federal Emergency Management Agency
CPR: Cardiorespiratory Resuscitation
CMO: Crew Medical Officer
MCC: Mission Control Centre
ACLS: Advanced Closed Loop System
ATLS: Advanced Trauma Life Support
ECG: Electrocardiogram
CT: Computed Tomograph
MRI: Magnetic resonance imaging
CHeCS: Crew Health Care Systems
SR: Situation Room
DSRM: Design Science Research Methodology
ESA: European Space Agency
EVA: Extravehicular Activity
FE: Flight Engineer
CMO: Crew Medical Officer
IV: Intravehicular
EV: Extravehicular
FD: Flight Director
FS: Flight Surgeon
BME: Biomedical Engineer
CAPCOM: Capsule Communicator
NASA-TLX: NASA’s Task Load Index
TEAM: Team Emergency Assessment Measure
PDAM: Pre-determined Debris Avoidance Maneuver
CRV: Crew Return Vehicle
NIHSS: NIH Stroke Scale
FAST: Face drooping, Arm weakness, Speech difficulty, and Time to call for medical help
HTTP: Hypertext Transfer Protocol
ID: Identification
CMRS: Crew Medical Restraint System
ICE: Isolated, Confined, and Extreme
SA: Situation Awareness

## Acknowledgements

The authors gratefully acknowledge the entire team of professionals from IDeaS Lab for their support in the demonstration, with special recognition to Eng. Ana Martins, Eng. Matilde Santos, Eng. Sónia Viegas, and Dr. Mariana Figueiras for their valuable contributions. The authors also extend sincere thanks to Dr. Adrianos Golemis for his insightful suggestions during the design and development phases.

The authors acknowledge Fundação para a Ciência e Tecnologia for its financial support via the project UIDB/00667/2020 (UNIDEMI). This work was supported by the project “e-Hospital4Future: Building future through an innovated and digital skilled hospital”, Grant EU4Health: 101101190. SFA was supported by eHospital4Future/EU4Health, ref. 205220. In both cases, the funding body provided financial support for the research, with no involvement in the analysis or writing of the article.

## Appendix A (Simulation’s Decisions)

### Decision 1

Role: FD

Decision 1 (9:50:00): During an EVA and after a blackout period, what should you do?

A. Through CAPCOM, check how things are going with the EVs during the fixation of the new component for the new radiation study.
B. Wait for the astronauts to report any issues.
C. Check if the communications are back on.
D. Check with the rest of the MCC about the scheduled work.

Role: CAPCOM

Decision 1 (10:00:00): Coming from a blackout period. What to do?

A. Check how are the things are going with the all astronauts during the fixation of the new component for the new radiation study.
B. Wait for the astronauts to report any issue.
C. Check if the communications are back on.
D. Check with the rest of the MCC the scheduled work.

Roles: COMMANDER (IV2), FE-2(IV1), FS, BME, EVA1, EVA2 Keep the normal activities going.

### Decision 2

Role: FD

Decision 2 (9:55:00): With the information provided by CAPCOM, what’s the next step?

A. Stop the activities in the EVA to understand the current situation
B. Continue with normal activities.
C. Check if the communications are back on.
D. Check with the rest of the MCC about the scheduled work.

Roles: FS, BME

Decision 2 (9:55:00): With the information provided by CAPCOM to the FS, what’s the next step?

A. Ask, through CAPCOM, for the flight director to order a to stop the activities in the EVA to perform a medical assessment
B. Continue with normal activities.
C. Check if the communications are back on.
D. Check with the rest of the MCC about the scheduled work.

Role: CAPCOM

Check how are the things are going with the all astronauts during the fixation of the new component for the new radiation study. Report the status back to the flight director and flight surgeon.

Role: EVA1

You are now starting the fixation of the new component of data collection to the station. Communicate what you’re doing only when asked by the CAPCOM

Role: EVA2

You are now on a different location than FE-1(EV1), checking on the status of other colection device. Report the positive status of this device only when asked by the CAPCOM.

Role: COMMANDER (IV2) + FE-2(IV1)

Report your positive status only when CAPCOM asks

### Decision 3

Role: EVA1

Decision 3 (10:00:00): 2 hour into EVA, sundendly you feel numbness on your right arm while performing a vigurous task with that arm. What should you do?

A. Communicate with CAPCOM to report what you fell.
B. Initiate return to the crew lock while continuing to communicate with MCC.
C. Check for suit malfunctions before making any further decisions.
D. Communicate with Commander in the station to report what you fell.

Roles: EVA2, COMMANDER (IV2), FE-2(IV1), FD, CAPCOM, FS, BME

Keep the normal activities going.

### Decision 4

Role: CAPCOM

Decision 4 (10:00:30): You are in communication with FE-1(EV1). With the information he provided, what should you do?

A. Communicate the situation to the flight director and transfer communication to the Flight Surgeon. A private consultation with EVA-1 is recommended.
B. Instruct FE-1(EV1) to initiate the return to the crew lock.
C. FE-1(EV1) should check for suit malfunctions before making any further decisions.
D. Communicate with the Commander at the station to report what FE-1(EV1) feels.

Role: EVA1

Report to CAPCOM the numbness in your right arm.

Roles: EVA2, COMMANDER (IV2), FE-2(IV1), FD, CAPCOM, FS, BME

Keep the normal activities going.

### Decision 5

Role: FS

Decision 5 (10:01:00): You’re now in a private consult with FE-1(EV1). After gathering more information about the situation. What do you think should be suggested?

A. Instruct FE-1(EV1) to stop his work for 15 minutes. Rest in the anatomical reference position to see if the symptoms disappear.
B. Instruct FE-1(EV1) to initiate the return to the crew lock.
C. Instruct FE-1(EV1) to stop his work and check for suit malfunctions.
D. Instruct FE-2(IV1) to suit up and go pick up FE-1(EV1)

Role: EVA1

You’re now in a private consultation with the flight surgeon. Meanwhile, you report that the numbness could be from the strenuous work you were performing with that arm.

Role: CAPCOM

Establish a private consultation between FE-1(EV1) and the flight surgeon. Report the situation to the flight director and the IV astronauts.

Roles: EVA2, COMMANDER (IV2), FE-2(IV1), FD, BME

Keep the normal activities going.

### Decision 6

Role: FS

Decision 6 (10:16:00): With the information reported by FE-1(EV1). What is the best option?

A. You should report the situation to CAPCOM as a possible medical emergency.
B. Wait to see if the symptoms go away.
C. You should report the situation to CAPCOM and proceed with normal activities.
D. You should report the situation to BME.

Role: EVA1

After 15 min, the symptoms worsen, now you notice weakness in your right arm. You’re still in the private consultation with the fligh surgeon please report it.

Role: BME

Monitor FE-1(EV1) vital signs.

Roles: EVA2, COMMANDER (IV2), FE-2(IV1), FD, CAPCOM, BME

Keep the normal activities going.

### Decision 7

Role: FD

Decision 7 (10:17:00): Receiving the information from the flight surgeon and knowing that the two EVAS aren’t next to each other. What should be done?

A. Instruct EVA2 to meet EVA1 to provide him with any assistance, even though this deslocation could take up to 20 minutes.
B. Instruct FE-3(EV2) to initiate the return to the crew lock.
C. Instruct FE-1(EV1) to initiate the return to the crew lock.
D. Ask another astronaut (still inside) to start preparing to come as EVA3 and pick EVA1

Roles: FS, BME

Decision 7 (10:17:00): Report the situation to the CAPCOM as a possible medical emergency. In discussion with BME/FS, what should be reported?

A. Confirm that this is a possible medical emergency and call for a trauma medical specialist. Meanwhile, BME monitors FE-1(EV1) vital signs.
B. Ask EVA1 to continue with the mission tasks and monitor the symptoms for the next 30 minutes.
C. Instruct EVA2 to take over EVA1’s tasks while EVA1 rests inside the airlock. No need to report yet unless the condition worsens.
D. Attribute the symptoms to suit fatigue and recommend hydration and deep breathing exercises before continuing. Meanwhile, BME monitors FE-1(EV1) vital signs.

Role: CAPCOM

Report the situation to the flight director

Role: FE-1(EV1)

Wait for more instructions.

Roles: COMMANDER (IV2), FE-2(IV1), FE-3(EV2)

Keep the normal activities going.

### Decision 8

Role: FS

Decision 8 (10:18:00): What can be done at this stage to help narrow down or confirm a diagnosis for EVA1’s condition?

A. Instruct FE-3(EV2) to perform the FAST method; Assess the 6 P’s (pain, pallor, pulselessness, perishingly cold, paraesthesia and paralysis); Monitor for headache severity and photophobia on FE-1(EV1)
B. Instruct FE-3(EV2) to assess orthostatic vital signs; Perform a neurological exam focused on awareness and responsiveness; Monitor for emotional triggers or psychological stressors; Rule out dehydration or hypoxia; on FE-1(EV1)
C. Instruct FE-3(EV2) to perform a basic cognitive and coordination test; Assess for confusion, gait disturbances, or eye movement issues; Check for nutritional deficiencies; Conduct vestibular function tests (if feasible in-mission) on FE-1(EV1)
D. Instruct FE-3(EV2) to repeat FAST over time to evaluate symptom progression; Observe for recurrence or resolution of symptoms; Inspect for isolated facial nerve involvement; Conduct environmental checks; Screen for history of migraines or visual disturbances on FE-1(EV1)

Role: CAPCOM

Instruct FE-3(EV2) to meet FE-1(EV1) to provide him with any assistance.

Role: FE-3(EV2), FE-1(EV1)

Wait for instructions.

Role: BME

Monitor FE-1(EV1) vital signs.

Roles: COMMANDER (IV2), FE-2(IV1), FD

Monitor the situation and assist with anything that the rest of the team might need.

### Decision 9

Role: CAPCOM

Decision 9 (10:38:00): You receive some more informationaboutf the situation from FE-3(EV2). What should you do?

A. Establish communication between the Flight Surgeon and FE-3(EV2) and report the situation to the flight director.
B. Ask EVA1 to perform basic speech and motor tests to confirm if it’s truly a neurological issue.
C. Instruct FE-3(EV2) to keep monitoring EVA1 for further symptoms and wait to see if the condition progresses.
D. Report the situation directly to the Flight Director and include the Flight Surgeon in the loop.

Role: FS

Decision 9 (10:38:00): Meanwhile, during communication, you notice that EVA1 is having difficulty speaking clearly. What should be done?

A. Communicate with FE-3(EV2) to understand what is happening. With this information it’s possible to understand the result of the FAST method.
B. Wait until EVA1’s speech improves to avoid unnecessary panic or false alarms.
C. Tell EVA1 to hydrate and rest briefly, then retry communication.
D. Ask CAPCOM to re-establish comms in case the distortion is due to a technical glitch.

Role: FD

Decision 9 (10:38:00): Receiving information from CAPCOM. In this situation, what’s the best option?

A. Abort EVA, instructing the EVs to return to the crew lock and the IVs to prepare the crew lock for repressurization. The return will take at least 15min.
B. Don’t abort the EVA yet. Wait to get a better understanding of the situation.
C. Instruct FE-3(EV2) to enter the airlock first to begin repressurization preparation while FE-1(EV1) follows at their own pace.
D. Initiate full depressurisation of the airlock while FE-1(EV1) is still en route to save time.

Role: FE-3(EV2)

After 20 minutes, you get close to FE-1(EV1) and notice that he is presenting some facial asymmetry, specifically drooping on the right side of the face. You perform the FAST method with positive results; Report this information to CAPCOM.

Role: BME

Monitor FE-1(EV1) vital signs.

Role: EVA1

You notice that you can’t feel the right part of you’re face, which makes it difficult to speak

Roles: COMMANDER (IV2), FE-2(IV1)

Monitor the situation and assist with anything that the rest of the team might need.

### Decision 10

Role: EVA2

Decision 10 (10:39:00): Receiving information from the CAPCOM, You are now escorting EVA1 back to the airlock. What is the most effective way to support EVA1 during this phase?

A. Maintain physical proximity, monitor EVA1’s mobility and responsiveness, and provide verbal reassurance while relaying updates to CAPCOM.
B. Minimize communication to reduce stress and interference with MCC telemetry.
C. Leave EVA1 briefly to speed up airlock prep steps from outside.
D. Focus on finishing any remaining mission objectives while EVA1 proceeds slowly toward the airlock.

Roles: FS, BME

Decision 10 (10:39:00): The medical team of the MCC discuss the possible diagnosis. What can be happening with EVA 1?

A. Stroke; Acute upper limb ischemia; subarachnoid hemorrhage; hypoglycemia; Meningitis/encephalitis
B. Syncope; Conversion disorder
C. Wernicke’s encephalopathy; Ménièrés disease
D. Transient ischemic attack (TIA); Bell’s palsy; Migraine with aura; Carbon monoxide exposure

Roles: COMMANDER (IV2), FE-2(IV1)

Decision 10 (10:39:00): Upon receiving the orders from CAPCOM, what technical procedures must be prioritised to ensure a safe and efficient re-entry to the airlock during this contingency?

A. Ensure suit telemetry is stable, initiate translation path clearance, verify airlock integrity, and coordinate repress sequence timing with MCC.
B. Switch EVA1’s suit to full manual mode to reduce data traffic and speed up the airlock cycle.
C. Instruct EVA2 to enter the airlock first to begin repressurization prep while EVA1 follows at their own pace.
D. Initiate full depressurisation of the airlock while EVA1 is still en route to save time.

Role: CAPCOM

Communicate the information provided by FD to FE-2(IV1)

Role: FD

Communicate with CAPCOM to abort EVA, instructing the EVs to return to the crew lock and the IVs to prepare the crew lock for repressurization.

Role: EVA1

Due to your simptoms you have difficulties communicating but you can still understand the situation.

### Decision 11

Role: EVA 2

Decision 11 (10:40:00): How should you transport FE-1(EV1) to the crew lock?

A. Tow EVA1 using a tether
B. Carry EVA1 using the SAFER unit
C. EVA1 should self-rescue using their propulsion system
D. Assist EVA1 into the MMSEV (Miniature Modular Space Exploration Vehicle)

Role: FS

Decision 11 (10:40:00): In these possible diagnosis, where should the medical assistance be performed?

A. Near the Airlock, where the Crew Medical Restraint System is possible to fixate;
B. Near where all the medical equipment is;
C. Outside the station
D. One needs to wait to leave the airlock

Role: EVA1

Due to your simptoms you have difficulties communicating but you can still understand the situation.

Role: BME

Monitor FE-1(EV1) vital signs.

Roles: COMMANDER (IV2) + FE-2(IV1) + FD + CAPCOM Keep preparing for the reentry of the EVs.

### Decision 12 (Key Decision)

Role: FD

Decision 12 (10:41:00): The repressurization of the crew lock can take up to 15 min. Given the circumstances, how should the pressurisation of the crew lock be conducted?

A. Partial pressurisation finishing at 12 psi (~10 min.)
B. Normal repressurization (~15 min.)
C. Emergency pressurization at a rate of 1.0 psi/second (~5 min)

### Decision 13

Role: FS

Decision 13 (10:42:00): To understand the situation, it’s important to establish the last known normal (the last time when EVA1 was well, not presenting symptoms). What do you think it is?

A. 10:16:00 from then, they have 60 minutes to administer the medication to have the best possible outcome.
B. 10:50:00 from then, they have 20 minutes to administer the medication to have the best possible outcome.
C. 11:16:00 from then, they have 30 minutes to administer the medication to have the best possible outcome.
D. 11:02:00 from then, they have 50 minutes to administer the medication to have the best possible outcome

Roles: COMMANDER (IV2), FE-2(IV1)

Decision 13 (10:42:00): While the EVs are returning, what should you (IVs) do to prepare for their arrival?

A. The Commander can confirm the medical emergency protocols with the Flight Surgeon; Flight Engineer 2 can get the medical equipment needed and the instructions on how to adjust airlock repressurization speed
B. The commander and Flight Engineer 2 can initiate a full cabin depressurisation to speed up EVA2’s reentry
C. The Flight Engineer 2 can leave the airlock hatch open and wait to pull the patient in manually; the Commander gathers information about the medical emergency
D. The crew can begin unrelated maintenance tasks to stay on schedule

Role: FD

Communicate with CAPCOM the type of repressurization that will be performed.

Role: CAPCOM

Report the information given by the FD to the astronauts

Role: EVA1

Due to your simptoms you’re no longer able to participate in any discussion

Role: EVA2

If asked the last known normal of the FE-1(EV1) state was 10:16:00

Role: BME

Monitor FE-1(EV1) vital signs.

### Decision 14

Role: FE-2(IV1)

Decision 14 (10:45:00): To prepare for the EVs arrival FE-2(IV1) will need to go grab medical kits. Discuss with flight surgeon what medical kits you should bring (maxim. 5).

A. Crew Medical Restraint System (CMR)
B. Emergency Medical Treatment Pack - Medications (Red) & Other components
C. Medical Diagnostic Pack (Blue);
D. Medical Supply Pack (Green);
E. Minor Treatment Pack (Pink);
F. Oral Medication Pack (Purple);
G. Physician Equipment Pack (Yellow);
H. Topical & Injectable Medication Pack - Medications (Brown);
I. Convenience Medication Pack (White);
J. IV Supply Pack (Gray)
K. Advanced Life Support Pack (ALSP) & Other Componentes

Role: FE-3(EV2)

You’re almost arriving to the Airlock.

Role: EVA1

Due to your simptoms you’re no longer able to participate in any discussion.

Role: BME

Monitor FE-1(EV1) vital signs.

Role: FS

Give assistance to the IVs inside the station

Roles: COMMANDER (IV2), FD, CAPCOM

Start preparing for the crew lock for repressurization

### Decision 15 (Key Decision)

Role: FD

Decision 15 (10:46:00): EVs arrive at the crew lock. Due to the high-stress environment, it’s crucial that CAPCOM reminds EVAs of an important step in their training.

A. Instruct CAPCOM to remind the EVs to breathe frequently, do not hold respiration.
B. Instruct CAPCOM to remind the EVs to pay attention to the temperature of the Airlock
C. Instruct CAPCOM to remind the EVs to make sure the door is well closed
D. Instruct CAPCOM to remind the BME to keep monitoring EV1 vital signals

### Decision 17

Roles: COMMANDER (IV2), FE-2(IV1)

Decision 17 (10:51:00): Repressurization is now completed. Which sequence of actions must be completed before the hatch can be safely opened?

A. Verify that both the lock and the station cabin have equalized pressures, do leak and contamination checks to ensure safety, ensure crew readiness through verbal confirmation;
B. Wait for a verbal cue from any crew member, then open the hatch immediately since previous repressurization was successful, regardless of current sensor readings.
C. Rely solely on the environmental control system’s automated signal that pressures are equalized, ignoring any crew reports or backup sensor data, and open the hatch once that signal is received.
D. Open the hatch once one side shows a nominal 12 psi reading, even if the opposite side has not yet confirmed equal pressure, but continue to monitor the telemetry post-opening.

Role: CAPCOM

Report the information given by the IVs

Role: FD

Give support to IVs according to the information transmited by CAPCOM

Role: EVA1

Due to your simptoms you’re no longer able to participate in any discussion

Role: EVA2

The repressurization is now completed wait for the hatch.

Roles: BME, FS

Monitor FE-1(EV1) vital signs.

### Decision 18

Roles: COMMANDER (IV2), FE-2(IV1)

Decision 18 (10:52:00): You proceed with the opening of the hatch. What are you doing first?

A. Doff the EVA1 suit.
B. Doff the EVs suits at the same time.
C. Doff the EVA 2 suit
D. Give proper medication to EVA 1

Roles: FS, BME

Decision 18 (10:52:00): What coordinated assessment could you do with the FS/BME perform during hatch opening and suit doffing?

A. Wait to see the vital signs data.
B. Rely solely on the telemetry data provided by the suit sensors, assuming that clinical symptoms will resolve once environmental pressures are equalised.
C. Conduct a evaluation of the telemetry data displayed.
D. Instruct EVA1 to self-monitor his condition while the BME temporarily disables any conflicting sensor readings to avoid false alarms.

Role: CAPCOM

Report the status of the station to the rest of the MCC

Role: FD

Give support to IVs according to the information transmited by CAPCOM

Role: EVA1

Due to your simptoms you’re no longer able to participate in any discussion

Role: EVA2

You’re still inside the EVA suit. This way, you need to wait for help to doff the suit.

### Decision 19

Roles: FS, BME

Decision 19 (11:01:00): In discussion with FS/BME and without any telemetry data, which immediate action should be prioritised right after the doffing of FE-1(EV1)? Communicate your decision to the MCC

A. Instruct IVs to perform a basic vital signs check (heart rate, blood pressure, and oxygen saturation), then resume concurrent tasks.
B. Instruct IVs to restrain FE-1(EV1) to the Crew Medical Restraint System and measure vital signs, perform neurological evaluation, while also checking for temperature regulation issues.
C. Instruct IVs to delay the assessment to avoid disrupting mission timelines and opt only to observe visual cues of distress, while keeping him in the current unsuited position.
D. Instruct IVs to quickly re-suit EVA 1 without assessment and plan for a full evaluation once his suit is secured.

Role: EVA1

Due to your symptoms you’re no longer able to participate in any discussion

Role: EVA2

You’re still inside the EVA suit. This way, you need to wait for help to doff the suit.

Role: CAPCOM

Report the decision of the FS to the IVs and FD

Roles: COMMANDER (IV2), FE-2(IV1), FD

Wait for more information.

### Decision 20

Role: FD

Decision 20(11:06:00): If the PDAM isn’t possible, what is the best option for the astronauts’ crew?

A. The astronauts crew start preparing for possible isolation in CRV (Crew Return Vehicle)
B. The astronauts crew remain in the current location.
C. The astronauts crew delay any further action while monitoring the PDAM to see if it eventually resumes functioning.
D. The astronauts crew initiate a spacewalk to attempt manual repairs on the PDAM while continuing other operations.

Role: FS

Decision 20(11:06:00): Given the information provided by the CAPCOM. What should you do first?

A. Relocate the site of treatment to the CRV
B. Wait for the patient in EVA 1 to improve with the ongoing treatment.
C. Delay any immediate actions and continue to monitor the patient’s condition in hopes that they stabilize.
D. Keep treating the patient in the current EVA 1 location without moving to the CRV, even though relocation may offer a safer or more controlled environment.

Role: BME

Monitor FE-1(EV1) vital signs.

Role: EVA1

Due to your simptoms you’re no longer able to participate in any discussion

Role: EVA2

You’re still inside the EVA suit. This way, you can’t move and need to wait for help to doff the suit.

Role: CAPCOM

Receive information from the MCC team and communicate with the astronauts.

Roles: COMMANDER (IV2), FE-2(IV1)

You are starting the restrainment of FE-1(EV1) to the Crew Medical Restraint System and measure vital signs of FE-1(EV1)

### Decision 21

Role: FS

Decision 21 (11:07:00): For the realocation of the site, what is the most appropriate action to take regarding EVA 2?

A. Suspend all further unsuiting activities for EVA 2 until the Flight Surgeon confirms that EVA 1’s physiological condition is stable.
B. Continue the unsuiting of EVA 2 currently with the ongoing primary assessment of EVA 1, ensuring both processes move forward in parallel.
C. Delay the primary assessment of EVA 1 until EVA 2 is fully unsuited, then evaluate both astronauts together.
D. Accelerate re-suiting procedures for EVA 1 by reallocating EVA 2’s timeline, so that EVA 1 can be quickly secured and evaluated.

Role: FD

Communicate the decision of Start preparing for possible isolation in CRV (Crew Return Vehicle) to CAPCOM

Role: CAPCOM

Communicate FD orders to the astronauts

Role: BME

Monitor FE-1(EV1) vital signs.

Role: EVA1

Due to your simptoms you’re no longer able to participate in any discussion

Role: EVA2

You’re still inside the EVA suit. This way, you can’t move and need to wait for help to doff the suit.

Roles: COMMANDER (IV2) + FE-2(IV1)

You start preparing to realocat the site of treatment to the CRV

### Decision 23

Role: FS

Decision 23 (11:16:00): Given the vital signs available. Which additional measurement should be obtained?

A. While FE-2(IV1) is doffing FE-3(EV2) from the EVA, Commander can measure the point-of-care blood glucose, serum electrolytes and perform NIH Stroke Scale
B. While FE-2(IV1) is doffing FE-3(EV2) from the EVA, Commander can measure the urinalysis and take a x-ray with portable chest radiograph
C. While FE-2(IV1) is doffing FE-3(EV2) from the EVA, Commander can do a toxicology screen and lumbar puncture
D. While FE-2(IV1) is doffing FE-3(EV2) from the EVA, Commander can do a serum amylase and lipase levels

Role: FE-2(IV1)

You’re now unsuiting EVA2

Role: COMMANDER (IV2)

You’re now assessing EVA 1 status

Role: FD

Communicate the decision of doff FE-3(EV2) suit to be able to go from the Quest Joint Airlock to the CRV (crew return vehicle)

Role: CAPCOM

Communicate FD orders to the astronauts, FS and BME

Role: EVA1

Due to your simptoms you’re no longer able to participate in any discussion

Role: EVA2

You’re now being unsuited from the EVA

Role: BME

Monitor FE-1(EV1) vital signs.

### Decision 24

Roles: FE-3(EV2), FE-2(IV1)

Decision 24(11:26:00): After completing the tasks communicated by the CAPCOM. You now start the relocation to the CRV. In discution with FE-3(EV2)/FE-2, how are you transporting FE-1(EV1)?

A. Attach EVA 1 securely to the Crew Mobility Restraint (CMR) system to minimise movement and maintain stability throughout transit.
B. Manually guide EVA 1 to the Crew Rescue Vehicle (CRV), ensuring controlled progress along a predetermined path.
C. Integrate both systems—first restrain EVA 1 using the CMR, then guide him toward the CRV—to maximise safety by merging immobilisation with controlled movement.
D. Allow EVA 1 to use a SAFER (Simplified Aid for EVA Rescue) unit for self-mobility while remaining tethered to a guideline, providing flexibility in movement coupled with a safety backup.

Role: FS

Decision 24(11:26:00): With the results of the point-of-care blood glucose, serum electrolytes and perform NIH Stroke Scale, what medical equipment do astronauts need to transport inside the CRV (max. 5)?

A. Crew Medical Restraint System
B. Emergency Medical Treatment Pack - Medications (Red) & Other components
C. Medical Diagnostic Pack (Blue);
D. Medical Supply Pack (Green);
E. Minor Treatment Pack (Pink);
F. Oral Medication Pack (Purple);
G. Physician Equipment Pack (Yellow);
H. Topical & Injectable Medication Pack - MediGreyons (Brown);
I. Convenience Medication Pack (White);
J. IV Supply Pack (Gray)
K. Advanced Life Support Pack (ALSP)

Role: FD

Receive information from CAPCOM

Role: CAPCOM

Communicate FS orders to Commander and FD

Role: BME

Monitor FE-1(EV1) vital signs.

Role: EVA1

Due to your simptoms you’re no longer able to participate in any discussion

Role: Commander

You’re in communication with MCC

### Decision 25

Roles: FE-3(EV2) + FE-2(IV1)

Decision 25(11:31:00): You and EVA1 are now inside the CRV. Which of the following sets of actions is required to properly prepare the Crew Rescue Vehicle (CRV) for a possible isolation scenario?

A. Close the hatches, don IV spacesuits, prepare CRV communications and systems
B. Open all external hatches to equalise pressure and bypass internal communications
C. Activate emergency thrusters, disengage life-support controls, and initiate a full system shutdown
D. Seal off the cabin ventilation, disable navigation systems, and switch to manual engine overrides

Role: FD

Receive information from the CAPCOM and assist in the decisions

Role: Commander

You perform the orders communicated by CAPCOM

Role: EVA1

Due to your simptoms you’re no longer able to participate in any discussion

Role: CAPCOM

Communicate the orders to the astronauts and FD

Role: BME

Monitor FE-1(EV1) vital signs.

Role: FS

Instruct, through CAPCOM, that the astronauts need to take the following medical kits inside CRV:

B. Emergency Medical Treatment Pack - Medications (Red) & Other components
F. Oral Medication Pack (Purple);
G. Physician Equipment Pack (Yellow);
K. Advanced Life Support Pack (ALSP)

### Decision 26

Role: FD

Decision 26(11:36:00): All astronauts are now inside the CRV. You’re in a joint communication between the flight director, CAPCOM and the astronauts. What’s the next step?

A. Do not initiate undocking in advance. Instead, the crew should wait until the moment of possible impact to see the situation. The crew should react based on the extent and location of any atmospheric loss.
B. Preemptively undock the CRV as soon as there is any indication of a potential collision, regardless of whether atmospheric loss is expected, to ensure maximum separation between the station and the spacecraft.
C. Delay all isolation procedures until the collision event is inevitable, then rapidly undock and separate the modules immediately after impact.
D. Immediately close all hatches, isolate the modules, and initiate undocking simultaneously without waiting for further analysis of the collision specifics. This ensures that the crew is separated as quickly as possible, even if it may result in unnecessary evacuation.

Roles: FS, BME

Decision 25(11:36:00): The medical team of the MCC. According to the results of the exams, what is the most likely diagnosis?

A. Ischemic Stroke
B. Hemorrhagic stroke
C. Hypoglycemia
D. It isn’t possible to do a diagnosis due to the lack of clinical information

Role:EVA1

Due to your symptoms you’re no longer able to participate in any discussion

Roles: CAPCOM, COMMANDER (IV2), FE-2(IV1), FE-3(EV2) You’re in a joint communication between the flight director, CAPCOM and the astronauts the possible collision situation

### Decision 27

Role: FD

Decision 27 (11:45:00): We have new information about the location of the object, and a collision is expected whithin 20min. What’s the correct order of tasks to isolate the crew inside the CRV?

A. Establish CRV communications with MCC, close the hatches, verify opportunity windows for undocking, check propulsion systems, perform delta-V calculations for orbital manoeuvring, assess propellant requirements (if necessary), and don the IV spacesuits.
B. Establish CRV communications with MCC, check propulsion systems, close the hatches, perform delta-V calculations for orbital manoeuvring, verify opportunity windows for undocking, assess propellant requirements (if necessary), and don the IV spacesuits.
C. Verify opportunity windows for undocking, close the hatches, check propulsion systems, establish CRV communications with MCC, perform delta-V calculations for orbital manoeuvring, don the IV spacesuits, and assess propellant requirements (if necessary). D.Don the IV spacesuits, assess propellant requirements (if necessary), perform delta-V calculations for orbital manoeuvring, check propulsion systems, establish CRV communications with MCC, verify opportunity windows for undocking, and finally close the hatches.

Roles: FS, BME

Decision 27 (11:45:00): The medical team of the MCC. What’s the best course of treatment with the tools available?

A. Administer 325 mg aspirin orally
B. Initiate antihypertensive infusion to lower SBP toward < 140 mmHg
C. Check capillary blood glucose and correct any hypoglycemia
D. It isn’t possible to chose a treatment due to the fact that there’s not enough information to do a diagnosis

Role: EVA1

Due to your symptoms you’re no longer able to participate in any discussion

Roles: CAPCOM, COMMANDER (IV2), FE-2(IV1), FE-3(EV2) You’re in a joint communication between the flight director, CAPCOM and the astronauts the possible collision situation

## Notes

### Competing Interest Statement

The authors have declared no competing interest.

### Funding Statement

The European Space Agency and the Portuguese Space Agency only funded the presentation of this work in the 76th International Astronautical Congress (IAC 2025)

